# BLM overexpression as a predictive biomarker for CHK1 inhibitor response in PARP inhibitor–resistant BRCA-mutant ovarian cancer

**DOI:** 10.1101/2022.12.02.22283037

**Authors:** Nitasha Gupta, Tzu-Ting Huang, Jayakumar R. Nair, Daniel An, Grant Zurcher, Erika J. Lampert, Ann McCoy, Ashley Cimino-Mathews, Elizabeth M. Swisher, Marc R. Radke, Christina M Lockwood, Jonathan B. Reichel, Chih-Yuan Chiang, Kelli M. Wilson, Ken Chih-Chien Cheng, Darryl Nousome, Jung-Min Lee

**Affiliations:** Women’s Malignancies Branch, Center for Cancer Research (CCR), National Cancer Institute (NCI), National Institutes of Health (NIH), Bethesda, Maryland, 20892, USA; Department of Obstetrics and Gynecology, Cleveland Clinic, Cleveland, Ohio, 44195, USA; Departments of Pathology and Oncology, Johns Hopkins University School of Medicine; Baltimore, Maryland, 21231-2410, USA; Brotman Baty Institute of Precision Medicine, University of Washington, Seattle, Washington, 98195, USA; Department of Laboratory Medicine and Pathology, University of Washington, Seattle, Washington, 98195, USA; National Center for Advancing Translational Sciences, NIH, Rockville, Maryland, 20892-4874, USA; Center for Cancer Research Collaborative Bioinformatics Resource, CCR, NCI, NIH, Bethesda, Maryland, 20892, USA

## Abstract

PARP inhibitors (PARPis) have changed the treatment paradigm in *BRCA*-mutant high-grade serous ovarian carcinoma (HGSC). However, most patients eventually develop resistance to PARPis, highlighting an unmet need for novel therapeutic strategies. Using high-throughput drug screens, we identified ATR/CHK1 pathway inhibitors as cytotoxic, and further validated monotherapy activity of the CHK1 inhibitor (CHK1i), prexasertib, in PARPi-resistant *BRCA*-mutant HGSC cells and animal models. As a proof-of-concept trial, we conducted a phase II study of prexasertib in *BRCA*-mutant HGSC patients. The treatment was well-tolerated but yielded an objective response rate of 6% (1/17; 1 PR) in patients with prior PARPi treatment. Exploratory biomarker analyses revealed that replication stress and fork stabilization were associated with clinical benefit to CHK1i. In particular, overexpression of *BLM*, and *CCNE1* overexpression or copy number gain/amplification were seen in patients deriving durable benefit from CHK1i. Our findings suggest replication fork–related biomarkers should be further evaluated for CHK1i sensitivity in HGSC.

**One Sentence Summary:** Overexpression of RecQ helicase BLM is a predictive biomarker for CHK1 inhibitor response in PARP inhibitor–resistant *BRCA*-mutant ovarian cancer.

## INTRODUCTION

High-grade serous ovarian carcinoma (HGSC), a lethal form of ovarian cancer, often presents at advanced stages, with 80% of patients experiencing recurrences after initial therapy (*1*). Nearly half of HGSCs exhibit deficiency in homologous recombination (HR) DNA repair due to defects in *BRCA1* or *BRCA2* and other HR-related genes, making them ideal candidates for poly (ADP ribose) polymerase inhibitor (PARPi)-based therapies (*2, 3*). Eventually, most patients discontinue PARPis due to progression, and the optimal management of PARPi-resistant HGSC is a pressing clinical challenge. To date, clinical studies have characterized a few mechanisms of resistance to PARPis that include restoration of HR repair by *BRCA* or *RAD51* reversion mutations, overexpression of *BRCA* hypomorphs, and other epigenetic means (*4*). However, very little is known about other resistance mechanisms prevalent in PARPi-resistant HGSC, highlighting the importance of identifying biomarkers that can help predict response to novel therapies.

Among the known resistance pathways exploited by PARPi-resistant tumors (*5*), replication fork stabilization, an outcome of increased fork protection from degradation (*e*.*g*., loss of PTIP expression or loss of SLFN11) or cell cycle checkpoint activation (*4*), is a major cause of PARPi resistance. It has been extensively studied in cell line models, but its validity and applicability as a biomarker in the clinical setting remain elusive (*5*).

In addition, the ATR/CHK1 pathway has been investigated in relation to DNA replication and fork stabilization (*6*). For instance, ATR and CHK1 phosphorylate nucleases (*e*.*g*., MUS81 (*7*)) and helicases (*e*.*g*., SMARCAL1 (*8*) and MCM2-7 (*9*)), thereby suppressing their activities and limiting fork reversal for replication fork stabilization. Studies have also identified overexpression of the ATR/CHK1 pathway in PARPi-resistant or platinum-resistant HGSC (*10, 11*). Furthermore, HGSC has nearly universal *TP53* mutation (*2*), thus disrupting the G1/S cell cycle checkpoint and rendering it more dependent on the ATR/CHK1-mediated G2/M checkpoint for DNA repair, making ATR/CHK1 signaling an attractive treatment target in *BRCA*-mutant PARPi-resistant tumors (*10, 11*). However, earlier clinical trials using ATR/CHK1 pathway blockade as monotherapy have revealed only modest clinical activity (*6*). For example, berzosertib (M6620) monotherapy yielded a response rate of 6% in patients with advanced or recurrent solid tumors that progressed on PARPis (*12*), highlighting the need for predictive biomarkers.

In the current study, based on our preclinical findings using PARPi-resistant HGSC cells (*13*), we hypothesized that patients with PARPi-resistant *BRCA*-mutant HGSC would gain clinical benefit from CHK1 inhibitor (CHK1i) treatment, and that biomarkers associated with replication fork dynamics would predict this clinical benefit. We tested the clinical activity and safety of the CHK1i prexasertib in heavily pretreated patients with *BRCA*-mutant HGSC, most with acquired PARPi resistance (NCT02203513). Our correlative studies also examined potential biomarkers associated with replication fork stabilization, such as the RecQ helicase BLM, which is essential for DNA resection and replication fork protection (*14*). The results of the clinical trial as presented here are the first demonstration of replication fork–related biomarkers that may predict clinical benefit to CHK1i in PARPi-resistant *BRCA*-mutant HGSC.

## RESULTS

### High-throughput drug screening identifies cell cycle checkpoint inhibitors as active drugs in PARPi-resistant *BRCA*-mutant ovarian cancer cell lines

To identify drug candidates for PARPi-resistant HGSC, we first performed high-throughput single drug screening using the National Center for Advancing Translational Sciences (NCATS) mechanism interrogation plate (MIPE) 5.0 library of 2,450 compounds (*15*) in three *BRCA2*-mutant HGSC cell lines: acquired PARPi-resistant PEO1/OlaR (*16*) along with *its parental PEO1* (*BRCA2* mutation 5193C>G [Y1655X]) and *de novo* PARPi-resistant PEO4 (*BRCA2* reversion mutation 5193C>T [Y1655Y]) (Table S1). Among others, 1,082 oncology drugs, both approved and investigational, were prioritized based on their status in clinical trial development (Fig. 1A and table S1). Z-transformed area under the curve (Z-AUC) (*15*) was used to distinguish inactive and active drug responses. Drugs with average Z-AUC values less than -1.0 in all three cell lines were classified as ‘hits’, resulting in 151 oncology drug hits (Table S2). The screen also confirmed cross-resistance between olaparib and other PARPis (rucaparib, niraparib, and talazoparib) in PARPi-resistant PEO1/OlaR and PEO4 (Fig. 1A and table S1). Notably, drugs against cell cycle checkpoint pathways were most enriched in the hits (20.5%, 31/151), followed by those targeting DNA replication (15.2%, 23/151), tubulin modulation (13.2%, 20/151), the PI3K/AKT pathway (11.9%, 18/151), and DNA repair (6.0%, 9/151) (Fig. 1B and table S2). Among the cell cycle checkpoint pathway inhibitors, prexasertib was ranked in the top 15 across all oncology drug hits and demonstrated significant cytotoxicity in all cell lines (Rank 15, Table S2), and it was used for subsequent preclinical studies and a clinical trial as follows.

**Fig. 1.**
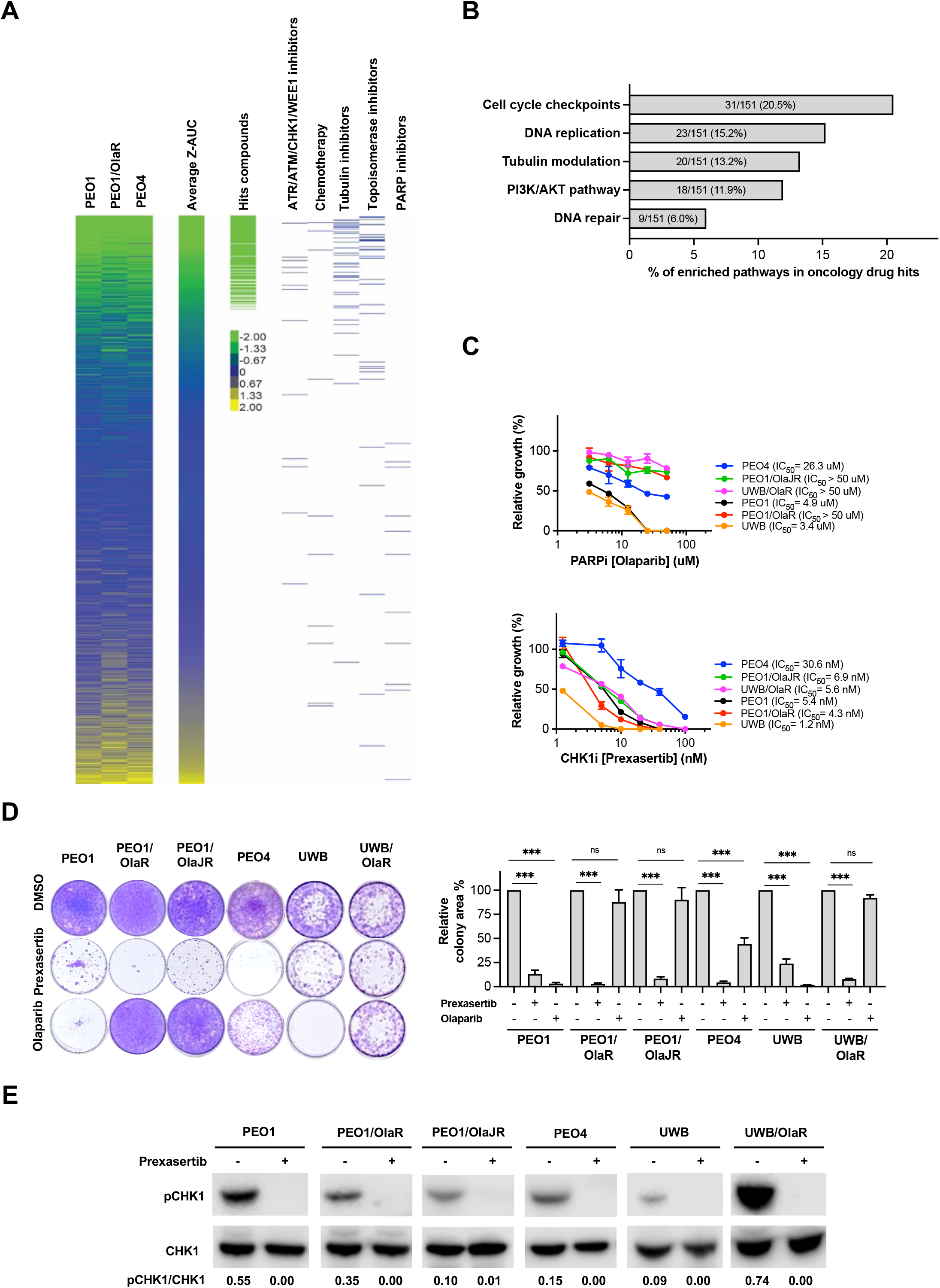
*In vitro* drug screen identifies ATR/CHK1 inhibitors as active drugs in both PARPi-sensitive and -resistant *BRCA*-mutant HGSC cells. (**A**) Heatmap of ranked drug activities for PARPi-sensitive and -resistant *BRCA2-*mutant HGSC cell lines including PARPi-sensitive PEO1, PARPi-resistant PEO1/OlaR, and *de novo* PARPi-resistant PEO4 cells. Single agent drug activities were screened using the MIPE 5.0 library of approved and investigational drugs (oncology drugs n = 1,082). Activity scores are based on Z-AUC. The average Z-AUC value > 0 indicates inactive drugs, while < 0 represents active drugs (*15*). The most active drugs based on their primary mechanisms of action, chemotherapy drugs for ovarian cancer, and PARPis are shown on the right. (**B**) Mechanistic classes enriched among highly active drugs are shown by Reactome database (https://reactome.org/) analysis. (**C-D**) Cell proliferation was validated using XTT (C) and colony-forming assays (D) in *BRCA1*-null UWB, *BRCA2*-mutant PEO1, *BRCA2*–reverse mutation PEO4, and acquired PARPi-resistant UWB1.289 (UWB/OlaR) and PEO1 (PEO1/OlaR and PEO1/OlaJR) cell lines. **(C)** Cells were treated with PARPi olaparib (top) or CHK1i prexasertib (bottom) at indicated doses for 72 hours. IC50 values were calculated using GraphPad Prism v7.1. (**D**) Long-term cell growth was examined using colony-forming assays. Cells were seeded at low density and treated with prexasertib (0.5 nM for UWB, 5 nM for all other cell lines) or olaparib (10 μM) and grown for 12–15 days. Colonies were visualized by 0.01% (w/v) crystal violet staining. Quantification was performed by ImageJ. (**E**) On-target effect of prexasertib was assessed by immunoblotting of p-CHK1 (S296) and total CHK1. Densitometric values of p-CHK1 (S296) relative to total CHK1 are shown. All experiments were repeated at least in triplicate and data are shown as mean ± SEM. ***, p < 0.001; ns, not significant. Abbreviations: CHK1i, CHK1 inhibitor; HGSC, high-grade serous ovarian cancer; PARPi, PARP inhibitor; UWB, UWB1.289; Z-AUC, Z-transformed area under the curve.

We further validated the findings from the drug screen using six *BRCA-*mutant HGSC cell lines, including four *BRCA2*-mutant (PEO1, PEO1/OlaR, and PEO4 as described above, plus another acquired PARPi-resistant PEO1/OlaJR (*13*) and two *BRCA1*-mutant (PARPi-sensitive UWB1.289 [UWB] and -resistant UWB/OlaR) cell lines. IC50 values against olaparib were at least 8-fold higher in PARPi-resistant *BRCA*-mutant cells, ranging from 26.3 to > 50 *μ*M relative to their parental cells (UWB1.289 and PEO1, IC50 3.4 and 4.9 *μ*M, respectively [Fig. 1C, top]). IC50 values for prexasertib monotherapy ranged from 1.2 nM to 30.6 nM in PARPi-sensitive and - resistant *BRCA*-mutant HGSC cells (Fig. 1C, bottom). Using clinically attainable concentrations of prexasertib (0.125 nM to 100 nM) (*17*), CHK1i alone significantly decreased the colony-forming ability in both PARPi-sensitive and PARPi-resistant cells (Fig. 1D), whereas the combination with olaparib did not yield synergism in all six HGSC cells (fig. S1A-B), consistent with a previous report in *BRCA1*-mutant PARPi-resistant HGSC patient-derived xenograft (PDX) models (*18*). Lastly, immunoblotting showed that prexasertib inhibited CHK1 activation (lower p-CHK1 S296) in all cell lines (Fig. 1E), indicating a target effect.

### CHK1 inhibition induces lethal replication stress and DNA damage in PARPi-resistant *BRCA*-mutant HGSC cells

CHK1 plays an important role in replication fork stabilization and HR repair (*6*). We therefore hypothesized that significant cytotoxicity of CHK1i monotherapy in PARPi-resistant HGSC cells would be associated with replication fork destabilization and impaired HR repair. To test this hypothesis, DNA fiber assays were performed by incubating cells with 5-Chloro-2’- deoxyuridine (CIdU) followed by 5-Iodo-2’-deoxyuridine (IdU), followed by treatment with or without CHK1i and PARPi (olaparib). A lower ratio of IdU/CldU, which suggests replication fork destabilization and hindered replication, was observed in both PARPi-resistant and PARPi-sensitive cells when treated with CHK1i alone (Fig. 2A). This was not further exacerbated by the addition of olaparib in all PARPi-resistant cell lines except PEO1/OlaJR and PEO4 (fig. S2A). To measure HR repair functionality, RAD51 foci were evaluated (*19*). Upon olaparib treatment, the number of cells with > 5 RAD51 foci significantly increased in all PARPi-resistant cells, attesting to HR restoration in these *BRCA*-mutant cells (Fig. 2B). As previously reported by us and others (*20, 21*), CHK1i abrogated olaparib-induced RAD51 foci in all cell lines (Fig. 2B and fig. S2B).

**Fig. 2.**
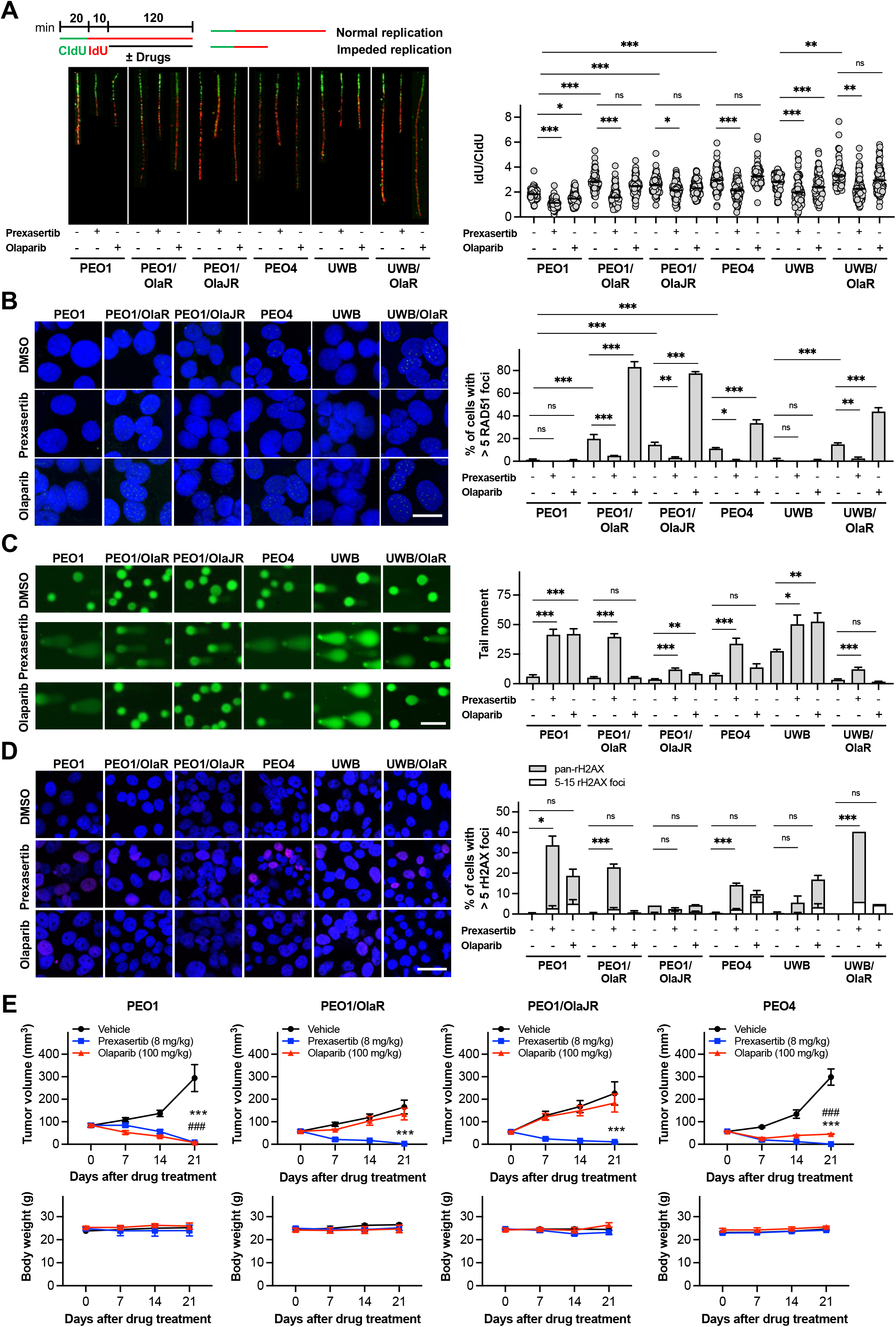
CHK1i monotherapy disrupts fork stabilization and HR restoration in PARPi-resistant *BRCA*-deficient HGSC models. (**A**) Replication fork stability in PARPi-sensitive (PEO1 and UWB), acquired PARPi-resistant (PEO1/OlaR, PEO1/OlaJR, and UWB/OlaR), and *de novo* PARPi-resistant (PEO4) cells were measured by DNA fiber assays. Cells were incubated with CldU, then IdU and co-incubated with prexasertib (20 nM) or olaparib (20 *μ*M). Representative images are shown (left). Dot plots of IdU (red) to CldU (green) tract length ratios in treated cells are plotted (right). A lower ratio of IdU/CldU indicates fork destabilization and hindered replication, suggesting higher replication stress. (**B-D**) Cells were treated with prexasertib (5 nM for all cell lines except 0.5 nM for UWB1.289 [UWB]) or olaparib (10 μM) for 48 hours. **(B)** HR status was measured by immunofluorescence staining of RAD51 foci. Representative images are shown (left) and the percentage of cells with > 5 RAD51 foci is plotted (right). Scale bar is 20 *μ*m. DNA damage was examined by (C) alkaline comet assay and (D) immunofluorescence staining of γH2AX foci (marker of DNA double-strand breaks). **(C)** Representative images of DNA fragments are shown (left) and the percentage of DNA in comet tails is plotted (right). Scale bar is 100 *μ*m. **(D)** The γH2AX foci (pink) were assessed by immunofluorescence staining. Cell nuclei were stained with DAPI (blue). Representative images were shown (left). The percentage of cells with 5–15 γH2AX foci, representing cells with DNA damage, and cells with pan-γH2AX staining, indicating cell apoptosis, is plotted (right). Scale bar is 50 μm. The above experiments were repeated at least in triplicate and data are shown as mean ± SEM. *, p < 0.05; **, p < 0.01; ***, p < 0.001; ns, not significant. (**E**) *In vivo* assessment of prexasertib (8 mg/kg) and olaparib (100 mg/kg) in PARPi-sensitive and PARPi-resistant *BRCA2*-mutant HGSC xenograft tumors (n = 5/group). Tumor volumes are plotted (top). The body weight of mice was measured once per week to monitor drug tolerance. No significant weight loss was found in mice with either prexasertib or olaparib treatment (bottom). Data are shown as mean ± SEM. ***, p < 0.001, prexasertib vs. vehicle; ###, p < 0.001, olaparib vs. vehicle. Abbreviations: CldU, 5-chloro-2′-deoxyuridine; CHK1i, CHK1 inhibitor; HR, homologous recombination; IdU, 5-Iodo-2′-deoxyuridine; PARPi, PARP inhibitor; UWB, UWB1.289.

DNA damage was assessed by alkaline comet assays, which showed increased comet tail moment with CHK1i monotherapy in all cell lines compared to the control (Fig. 2C). Immunofluorescent staining also showed increased γH2AX foci (DNA damage marker) and pan-γH2AX staining indicating apoptosis (*22*) in cells treated with CHK1i alone compared to the untreated group (Fig. 2D). Again, we did not observe any increase in either comet tail moment or γH2AX foci in PARPi-resistant cells when treated with CHK1i and olaparib compared to CHK1i alone (fig. S2C-D). In summary, our findings suggest that CHK1i as a monotherapy effectively overcomes PARPi resistance in *BRCA*-mutant HGSC by impairing replication fork dynamics and attenuating HR restoration, leading to DNA damage and lethal replication stress.

### CHK1 inhibition reduces tumor growth in PARPi-resistant *BRCA*-mutant HGSC murine models

To confirm the *in vitro* activity of CHK1i, we subcutaneously implanted *in vivo* models of immunodeficient NOD-SCID gamma (NSG) mice with PARPi-sensitive (PEO1) and PARPi-resistant (PEO1/OlaR, PEO1/OlaJR, and *de novo* PEO4) cells. CHK1i alone significantly reduced tumor growth in all murine models without notable weight loss (p < 0.001, Fig. 2E). In contrast, olaparib therapy had little effect on tumor growth in PEO1/OlaR and PEO1/OlaJR models and demonstrated some tumor growth inhibition in *de novo* PARPi-resistant PEO4 models, though not to the same extent as CHK1i (Fig. 2E). Together, these findings highlight the therapeutic potential of CHK1i in PARPi-resistant HGSC *in vivo*.

### CHK1 inhibition has modest antitumor activity and is well tolerated in PARPi-resistant HGSC patients with *BRCA* mutation

As our data indicated significant activity of the CHK1i prexasertib in PARPi-resistant *BRCA*-mutant HGSC *in vitro* and *in vivo* models, we conducted a proof-of-concept phase II clinical trial evaluating prexasertib activity in patients with *BRCA*-mutant recurrent HGSC (NCT02203513). Between February 2015 and July 2019, a total of 22 women were enrolled and received at least one dose of prexasertib (Fig. 3A and fig. S3A). The median age was 56.4 years (range, 35.7–74.8 years). Two-thirds of patients (15/22) had *BRCA1* mutation, and one-third (7/22) had *BRCA2* mutation. Most patients were heavily pretreated, with a median of 5 prior systemic treatments. Approximately 41% of patients (9/22) had platinum-sensitive disease and 59% (13/22) had secondary platinum-resistant disease. All patients had received platinum-based therapy and all except one had prior PARPis. The median duration of PARPi treatment was 9 months (range, 3.5– 48 months), and the median PARPi-free interval before starting the trial was 4.5 months (range, 1–26 months) (Table 1). All patients derived some degree of clinical benefit from prior PARPis, suggesting that PARPi resistance was acquired rather than *de novo* in this cohort.

**Table 1.**
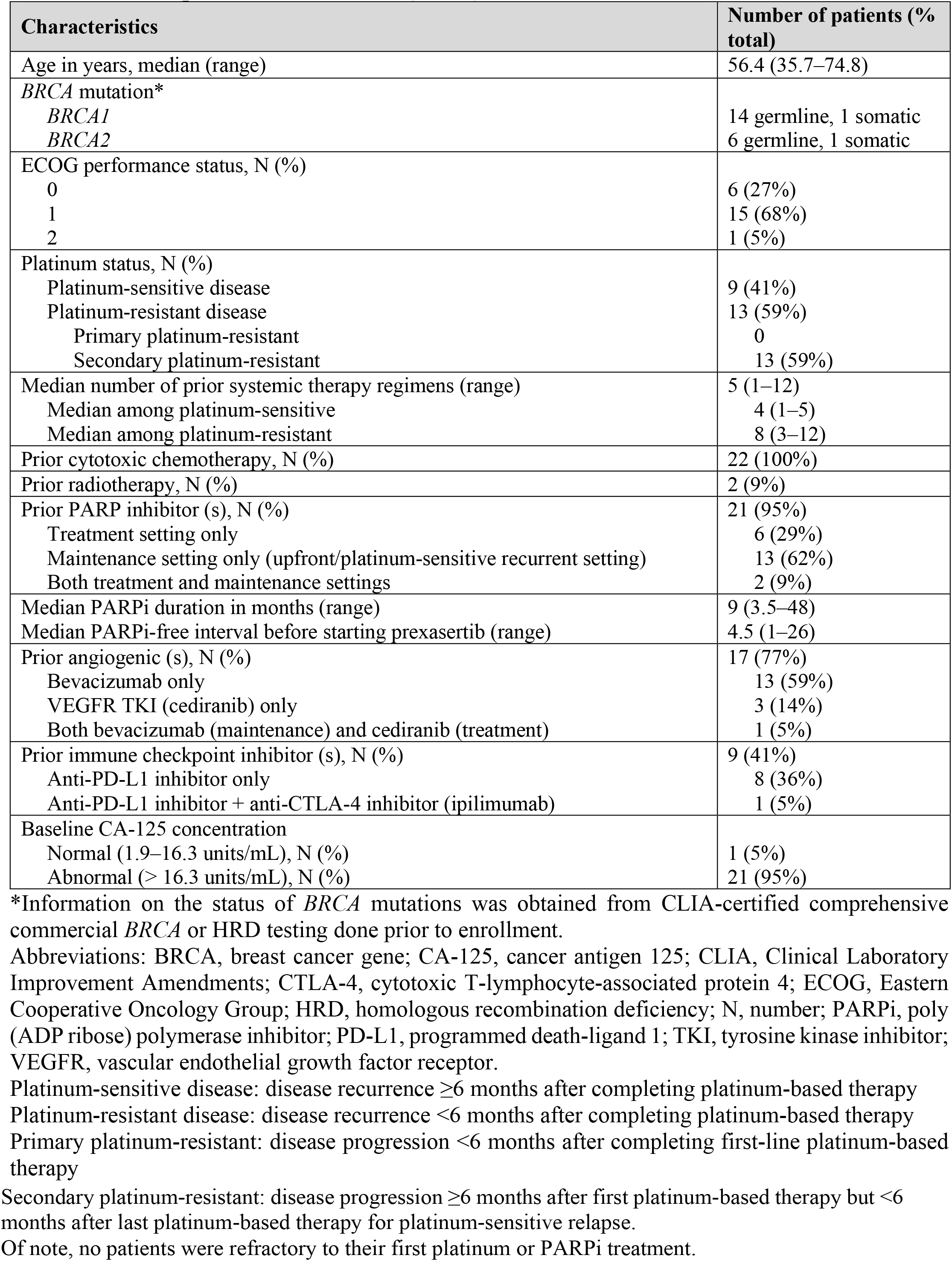
Baseline patient characteristics (n = 22)

**Fig. 3.**
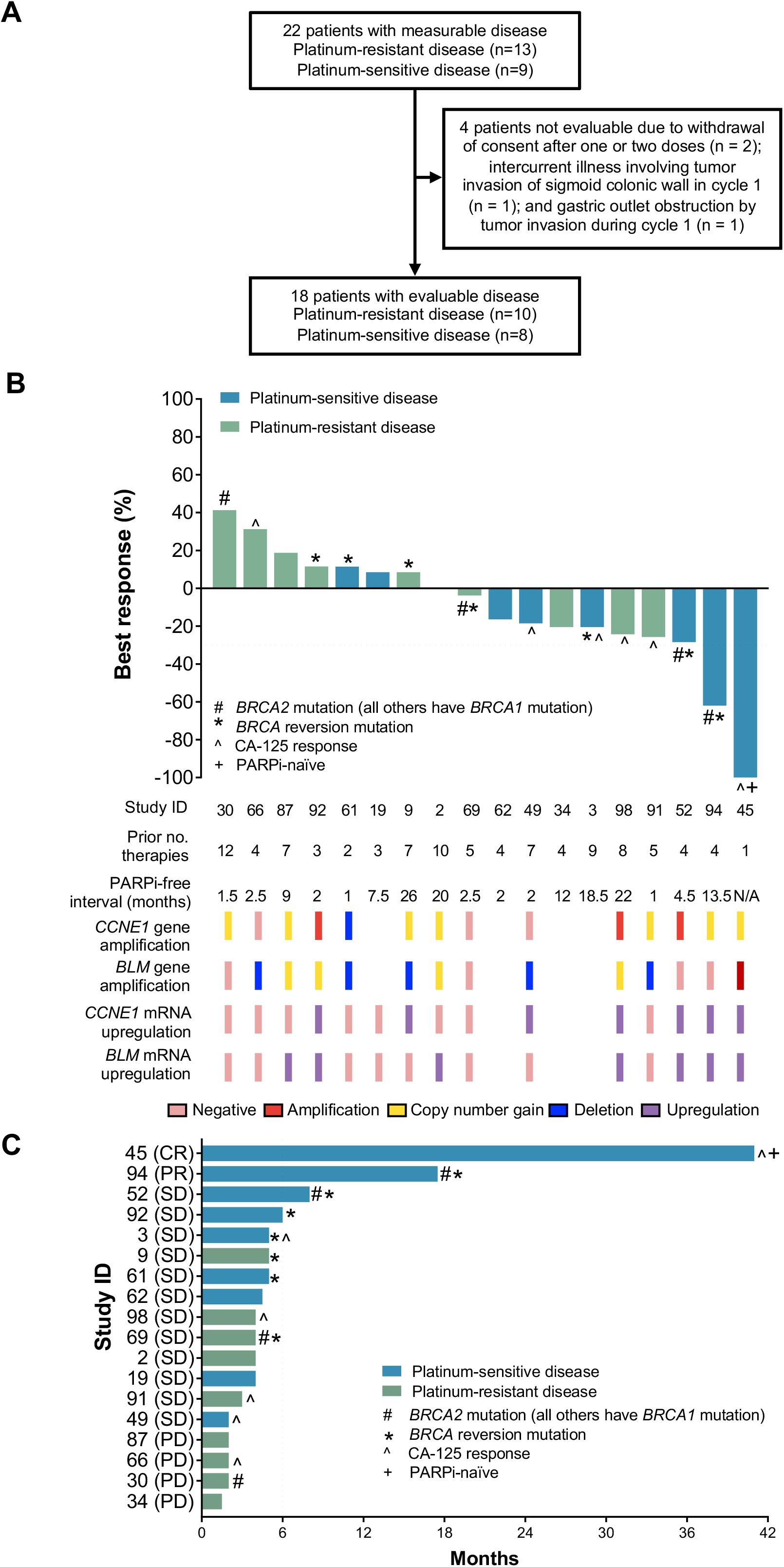
Clinical trial design and activity of prexasertib in BRCA-mutant HGSC. (**A**) The CONSORT flow diagram. (**B**) Waterfall plot: the best response to treatment in 18 evaluable patients. Best RECIST responses (percentage change from baseline in tumor size) are shown according to the number of previous treatments, PARPi-free interval, presence of *BLM* and *CCNE1* gene amplification/gain/deletion, and presence of *BLM* and *CCNE1* mRNA upregulation in pretreatment biopsy samples. The horizontal dotted line indicates the threshold for PR (30% reduction in tumor size from baseline). (**C**) Swimmer plot: duration of treatment of 18 evaluable patients. The vertical dotted line indicates the threshold for clinical benefit (CR + PR + SD ≥ 6 months). The cross symbol (+) in (B) and (C) indicates the PARPi-naïve, CR patient. The hash symbol (#) represents cancers with *BRCA2* mutation; all others have *BRCA1* mutations. Asterisks (*) indicate cancers with *BRCA* reversion mutations detected (see Table S4 for more details). The carrot symbol (^) indicates patients with CA-125 response (see fig. S3C for more details). Abbreviations: RECIST, Response Evaluation Criteria in Solid Tumors; CR, complete response; PR, partial response; SD, stable disease; PD, progression of disease. N/A, not applicable; no, number; PARPi, PARP inhibitor; R, platinum-resistant disease; S, platinum-sensitive disease.

Eighteen of 22 patients were evaluable for Response Evaluation Criteria in Solid Tumors (RECIST) v1.1 response (Fig. 3A). Of those, one attained complete response (CR), while another attained partial response (PR), yielding an objective response rate (ORR) of 11% (2/18) (Fig. 3B). The patient with CR (41 months of progression-free survival [PFS] and 39 months of duration of response) was the only PARPi-naïve patient in this study, thus resulting in an ORR of 6% (1/17) for previously PARPi-treated patients (Fig. 3B-C and fig. S3B). Twelve patients (12/18, 67%) had stable disease (SD) and four patients (4/18, 22%) achieved durable clinical benefit, defined as CR + PR + SD ≥ 6 months (Fig. 3C). There was no significant difference in clinical benefit by *BRCA1* versus *BRCA2* mutation (p = 0.26). Median PFS was 4 months (range, 1.5–41 months) among all 18 evaluable patients (Fig. 3C). Six (43%) of 14 evaluable patients had a Gynecologic Cancer Intergroup (GCIG) CA-125 response (50% reduction in CA-125 during treatment with confirmation after 4 weeks and pretreatment CA-125 greater than two times the upper limit of normal): 1 CR (41 months on study, with CA-125 response at 1 month of treatment); 4 SDs (ranging from 2–5 months on study, all with CA-125 response at 1 month of treatment), and 1 PD (2 months on study, with CA-125 response at 1 month of treatment) (Fig. 3B-C and fig. S3C). Four patients were not evaluable due to withdrawal of consent after one or two doses (two patients); intercurrent illness involving tumor invasion of sigmoid colonic wall in cycle 1 (one patient); and gastric outlet obstruction by tumor invasion during cycle 1 (one patient), making it too early to assess their response to the treatment.

All treated patients had at least one any grade treatment-emergent adverse event (AE) (Table 2 and Table S3). Prophylactic granulocyte colony-stimulating factor (G-CSF) was not routinely given on cycle 1 day 1 to check for nadir on cycle 1 day 8. Consistent with previous reports (*17, 23, 24*), the most frequently observed grade 3 or 4 toxicity was neutropenia on cycle 1 day 8 (18/22 patients [82%]), and only one patient developed febrile neutropenia. The nadir occurred approximately one week after each dose (median 8 days [range, 8**–**11 days]) and was transient ≤ 7 days (median 4 days [range, 2**–**13 days]), without necessitating dose reduction or discontinuation (data not shown). G-CSF was given for subsequent treatments to avoid treatment delays or dose reduction in most patients (16 of 18 patients) with grade 3 or 4 neutropenia on day 8 of cycle 1 (Table 2). No patients had dose reductions, and other non-hematologic AEs were relatively mild (Table S3). One death occurred during the study off-treatment follow-up period due to progression of disease (PD) in a patient whose best response was SD. In general, the toxicity profile mirrored that of the phase II cohort of patients with *BRCA*-wildtype HGSC (*24*). Overall, though prexasertib was largely well tolerated among a heavily pretreated population, its modest ORR, especially among patients with prior PARPis, prompted us to investigate possible cross-resistance between PARPis and CHK1i.

**Table 2.** Treatment-related adverse events.

| Adverse event | Maximum grade in all patients (n = 22) |  |  |
| --- | --- | --- | --- |
|  | 1–2 | 3 | 4 |
| Hematologic | Number of patients (% total) |  |  |
| Anemia | 21 (95%) | 1 (5%) | 0 |
| Neutropenia* | 1 (5%) | 3 (14%) | 15 (68%) |
| Leukopenia | 5 (23%) | 9 (41%) | 5 (23%) |
| Thrombocytopenia | 14 (64%) | 1 (5%) | 2 (9%) |
| Lymphopenia | 11 (50%) | 6 (27%) | 0 |
| Febrile neutropenia | 0 | 1 (5%) | 0 |
| Non-hematologic | Number of patients (% total) |  |  |
| Fatigue | 11 (50%) | 0 | 0 |
| Fever | 4 (18%) | 0 | 0 |
| Allergic reaction | 2 (9%) | 0 | 0 |
| Headache | 1 (5%) | 0 | 0 |
| Nausea | 11 (50%) | 1 (5%) | 0 |
| Vomiting | 5 (23%) | 1 (5%) | 0 |
| Rash (acneiform, maculopapular) | 4 (18%) | 0 | 0 |
| Diarrhea | 4 (18%) | 0 | 0 |
| Constipation | 3 (14%) | 0 | 0 |
| Abdominal Pain | 2 (9%) | 0 | 0 |
| Anorexia | 2 (9%) | 0 | 0 |
| Oral mucositis | 1 (5%) | 0 | 0 |

Patients could be counted under more than one preferred term.
\*16 of 18 (89%) patients with grade 3/4 neutropenia on cycle 1 day 8 received prophylactic growth factors for subsequent treatments to avoid treatment delay or dose reductions.

### *BRCA* reversion mutation does not confer resistance to CHK1 inhibition

We first questioned whether this limited clinical response to CHK1i in previously PARPi-treated *BRCA*-mutant patients correlated with *BRCA* reversion mutations, which have been associated with cross-resistance to PARPis and platinum drugs (*25, 26*). Pretreatment cell-free DNA (cfDNA) (n = 18) and paired tumor tissue samples were available in 15 patients for targeted next-generation sequencing to identify *BRCA* reversion mutations (Fig. 4A). *BRCA* reversion mutations were identified from cfDNA in 33% (6/18) of patients and from tissue samples in 33% (5/15) of patients (Table S4). Overall, *BRCA* reversion mutations did not appear to correlate with clinical outcome, as *BRCA* reversions were still seen in three patients with clinical benefit (two SDs [6 and 8 months of PFS] and one PR [17.5 months of PFS]). All six patients with *BRCA* reversion mutations in cfDNA had a best RECIST response of SD with PFS ranging from 4–8 months. Furthermore, all three evaluable patients with the best response of PD, who were resistant to CHK1i treatment at the first restaging scans, did not have *BRCA* reversion mutations. Of note, we previously reported a durable clinical benefit rate to prexasertib of 46% in *BRCA*-wildtype HGSC patients (*24*), suggesting that CHK1i demonstrates antitumor activity with functional BRCA. Taken together, these data suggest that *BRCA* reversion mutations are unlikely to confer cross-resistance to CHK1i therapy in PARPi-resistant tumors, although the findings are limited by the relatively small sample size.

**Fig. 4.**
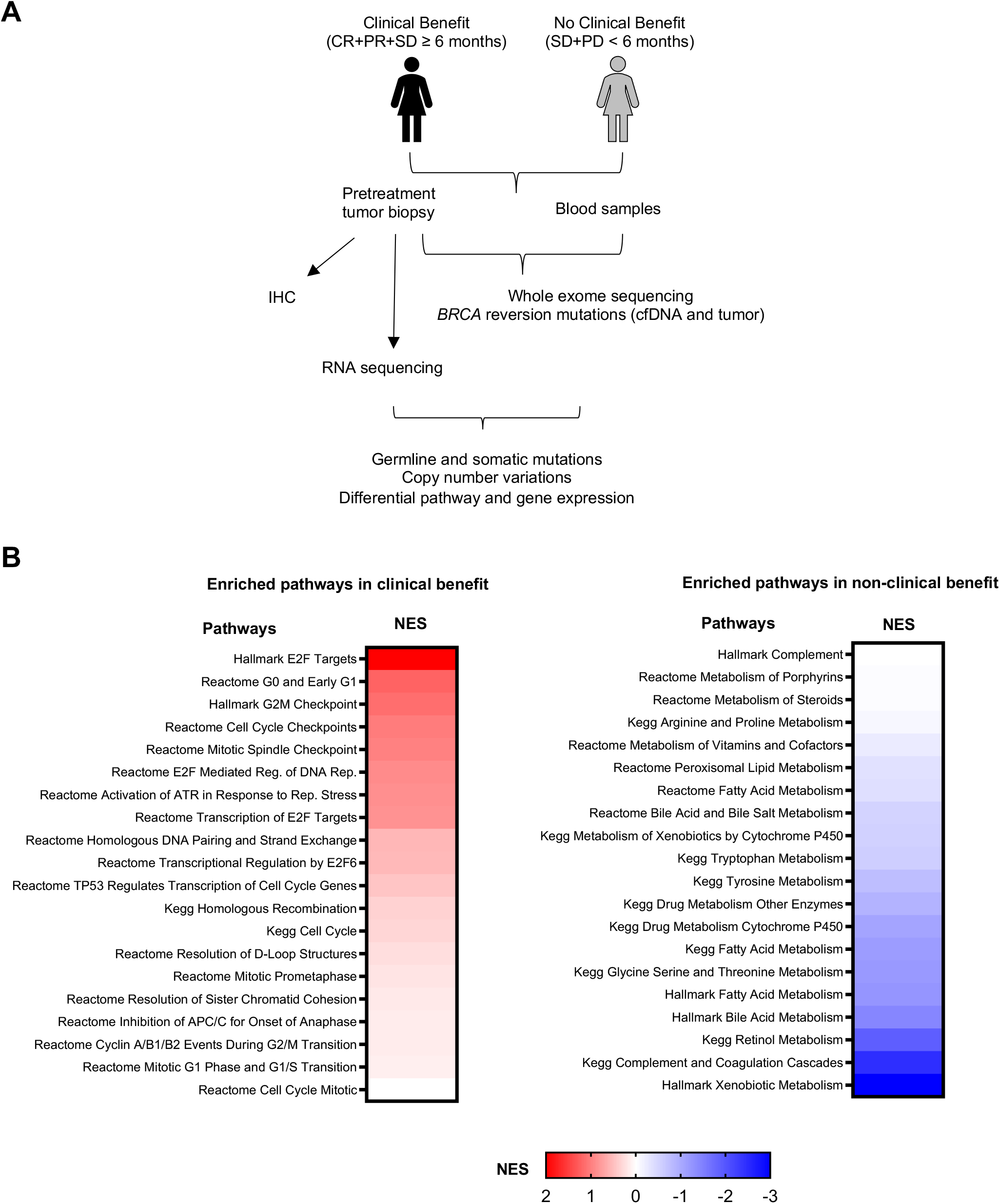
Exploratory biomarker analyses reveal differentially upregulated pathways with clinical benefit. (**A**) Schema for exploratory biomarker analyses. (**B**) Gene set enrichment analysis by Hallmark, KEGG, and Reactome databases of RNAseq show significantly upregulated (red) and downregulated (blue) gene set pathways in those with clinical benefit but not in non-clinical benefit group. Abbreviations: cfDNA, cell-free DNA; CR, complete response; IHC, immunohistochemistry; NES, normalized enrichment score; PD, progression of disease; PR, partial response; Rep, replication; SD, stable disease.

### Pathways related to high replication stress and low metabolism response are associated with clinical benefit to CHK1i in *BRCA*-mutant HGSC patients

Though sensitivity to CHK1i has been linked to decreased replication fork and DNA repair pathways in *BRCA*-wildtype HGSC cells and tumors (*21*), the pathways that can predict response to CHK1i in *BRCA*-mutant HGSC remain poorly understood. We thus performed transcriptomic analysis through RNA sequencing (RNAseq) on pretreatment biopsies obtained from patients deriving clinical benefit (CR + PR + SD ≥ 6 months, n = 4) versus those with no clinical benefit (SD + PD < 6 months, n = 11) to identify specific pathways related to CHK1i response (Fig. 4A and table S7). Gene set enrichment analysis (GSEA) using gene sets from the Hallmark, KEGG, and Reactome databases (*27*), demonstrated upregulation of pathways associated with cell cycle progression and DNA repair, including E2F target genes and the G2/M checkpoint, in the clinical benefit group relative to those with no clinical benefit, suggesting that patients deriving clinical benefit have tumors with high dependence on pathways to overcome replication stress (Fig. 4B, left). In contrast, those with no clinical benefit showed enrichment of pathways related to metabolism (Fig. 4B, right), which coincides with RNAseq findings on ATR inhibitors (ATRis) in cancers with high replication stress (*28*). These results also suggest that high levels of replication stress might be associated with clinical benefit to CHK1i, which is supported by what we had observed in PARPi-resistant *BRCA*-mutant cells (Fig. 2A).

### High levels of replication stress and replication fork stabilization are associated with clinical benefit to CHK1i in *BRCA*-mutant HGSC patients

Studies have shown that replication stress signatures that include genes related to HR and fork stability may predict the efficacy of ATR or CHK1 inhibitors in HGSC (*29, 30*), although their clinical utility, especially in the *BRCA*-mutant PARPi-resistant HGSC setting, requires further investigation. We asked whether the patients expressed specific genes that might indicate replicative stress and replication fork stabilization, and whether these genes correlated with response to CHK1i in *BRCA*-mutant PARPi-resistant tumors. Surprisingly, transcriptome profiling did not show any significant differences in the expression of genes associated with HR repair or PARPi resistance between the clinical benefit versus non-clinical benefit groups, although HR deficiency has been demonstrated to sensitize tumor cells to PARPis and DNA-damaging agents (*31*) (fig. S4 and table S8). In contrast, the expression of genes involved in replication stress and replication fork stability was increased in patients with clinical benefit (Fig. 5A and table S9). Therefore, we generated a list of 31 genes related to replication fork stabilization (*BLM, HLTF, FANCI, PARG, ZRANB3, TEN1* found in GSEA analysis) and known replication stress markers (*CCNE1, RB1, CDKN2A, KRAS, NF1, MYC, ERBB2, SRSF1, SUV39H1, GINS1, PRPS1, KPNA2, AURKB, TNPO2, ORC6, CCNA2, LIG3, MTF2, GADD45G, POLA1, POLD4, POLE4, RFC5, RMI1, RRM1*) in ovarian and other cancers (*29, 32*) to test which genes may be used as predictive biomarkers for CHK1i response. Notably, *BLM* and *CCNE1*, involved in fork stabilization and replication stress, were highly expressed in the clinical benefit group compared to the non-clinical-benefit group (Fig. 5B and table S9). Therefore, we hypothesized that tumors with high dependence on fork stabilization in response to replication stress, as indicated by increased *BLM* and *CCNE1* expression, would be more sensitive to CHK1i treatment in HGSC, and *BLM* could function as a potential biomarker for clinical benefit to CHK1i.

**Fig. 5.**
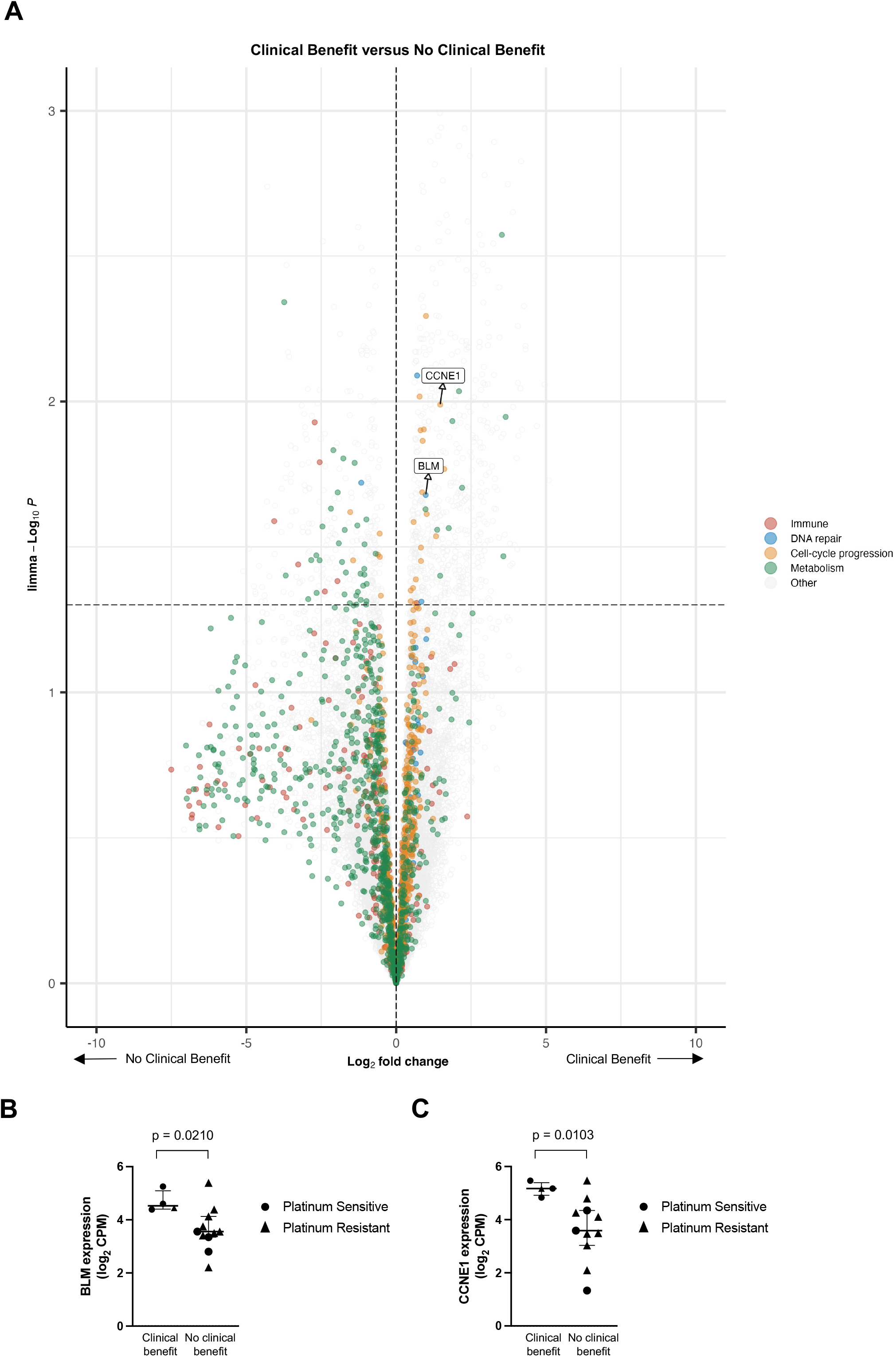
High mRNA expression of *BLM* and *CCNE1* are associated with clinical benefit. (**A**) The mRNA expression levels of genes were analyzed from RNAseq in patients with clinical benefit (n = 4) versus non-clinical benefit group (n = 11). The x-axis shows log2 fold change (>0 indicates genes enriched in patients with clinical benefit, while <0 indicates genes enriched in patients with no clinical benefit). The y-axis shows the Wilcoxon -log10 p values and the horizontal dotted line represents a significance threshold of p = 0.05, with significantly enriched genes falling above the line. (**B-C**) High mRNA expression of RecQ helicase *BLM* (B) and *CCNE1* (C) are significantly associated with clinical benefit. Lines represent median with 95% confidence interval. Abbreviations: CPM, counts per million.

Also, we analyzed their mRNA expression and prognostic impact using public Kaplan-Meier Plotter ovarian cancer database (*33*). No significant difference was found based on the expression of *BLM* (fig. S5A) or *CCNE1* alone (fig. S5B) but high/high co-expression of *BLM* and *CCNE1* in patients showed better PFS compared to those with high *BLM* and low *CCNE1* when treated with topotecan (median PFS 20.63 vs. 14.03 months, p = 0.045) (fig. S5C), supporting the notion that increased levels of *BLM* and *CCNE1* may serve as biomarkers of predicting the response of DNA replication inhibitors.

### BLM overexpression correlates with CHK1i sensitivity in PARPi-resistant *BRCA*-mutant HGSC cells

BLM is a member of RecQ helicase which is critical for replication fork stability by unwinding structures, *e*.*g*., G-quadruplex and Holiday Junctions for DNA replication (*34*). BLM also helps restart the stalled forks while suppressing firing of new origins in response to replication stress (*35*). We thus speculated that increased replication fork stabilization along with replication stress would better predict the sensitivity to CHK1i; furthermore, the role of BLM remains elusive in this phenomenon. To test this idea, we used PARPi-resistant *BRCA*-mutant HGSC cells to evaluate the role of BLM because of the paucity of clinical samples. We found that basal BLM levels were substantially higher in PARPi-resistant cells relative to their sensitive parental cells (Fig. 6A, top). Moreover, PARPi-resistant cells with high BLM levels showed markedly fewer (2.9–7.7%) surviving colonies when treated with CHK1i than their parental lines with lower BLM, demonstrating increased sensitivity of the former to CHK1i (Fig. 6A, bottom and fig. S6A). Further, the sensitivity of both BLM-low and BLM-high cell lines to CHK1i was enhanced when BLM was overexpressed in these cells (Fig. 6B). However, BLM overexpression induced sensitivity to CHK1i that was much greater (4.5–42-fold IC50) in PARPi-resistant cells than in their PARPi-sensitive parental cell lines (2.8 and 7.9-fold in PEO1 and UWB1.289 cells, respectively) (Fig. 6C). Cells overexpressing BLM also exhibited similarly enhanced sensitivity to another specific CHK1i, SRA737 (*36*) (fig. S6B). Taken together, these data suggest that high BLM expression in PARPi-resistant cells might predispose them to CHK1i treatment.

**Figure 6.**
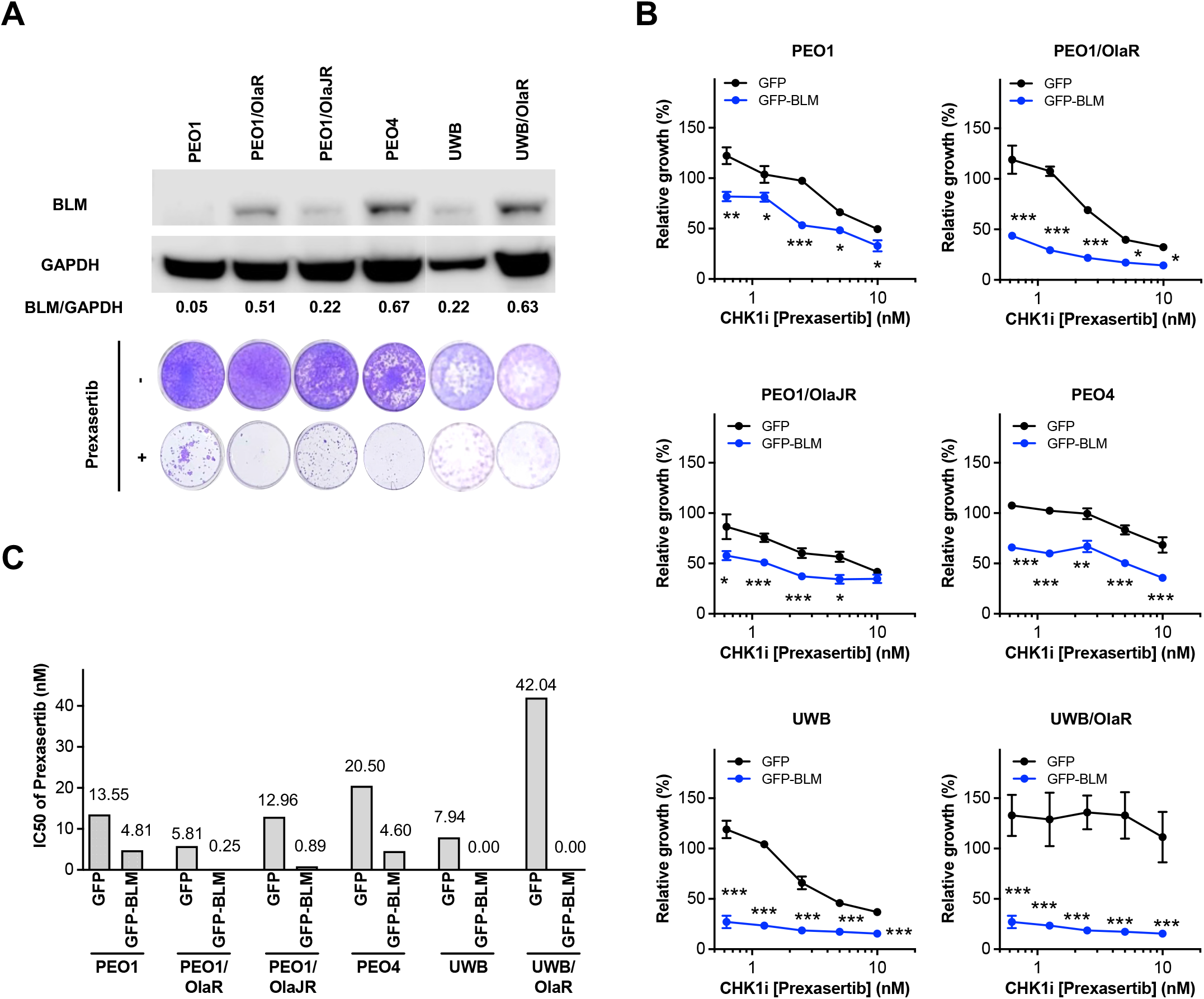
BLM overexpression increases the sensitivity of CHK1i in HGSC cell lines. (**A**) Basal levels of BLM in parental (*BRCA*-mutant PEO1 and UWB), acquired PARPi-resistant (PEO1/OlaR, PEO1/OlaJR, and UWB/OlaR), and *de novo* PARPi-resistant (PEO4) cell lines were analyzed by immunoblotting. Densitometric values of BLM relative to GAPDH (top) and representative colony formation images of cells with or without prexasertib treatment are shown (bottom). (**B-C**) Cells transfected with BLM overexpression plasmids for 48 hours were treated with or without prexasertib for another 48 hours. Cell viability was measured by XTT assay (B). IC50 values (C) from (B) were calculated using GraphPad Prism v7.1. Experiments were repeated at least in triplicate and data are shown as mean ± SEM. *, p < 0.05; **, p < 0.01; ***, p < 0.001. Abbreviations: CHK1i, CHK1 inhibitor; PARPi, PARP inhibitor; UWB, UWB1.289.

### Clinical benefit is associated with *CCNE1* amplification

In addition, we examined somatic variants in pretreatment tumors (n = 15) using whole-exome sequencing to identify genetic alterations associated with response to CHK1i. As expected, most tumors (13/15) had somatic mutations in *TP53*, consistent with previous reports (*2, 37*). Also, it has been hypothesized that HGSC with HR deficiency may present high tumor mutational burden (TMB) and benefit from drugs targeting DNA damage repair (*38*). However, in our cohort, TMB and somatic variants in HR-related genes were not significantly associated with clinical benefit (fig. S7 and table S5).

As reported previously (*39*), *CCNE1* amplification was seen in those with clinical benefit (Fig. S7). Seven (50%) of 14 evaluable patients had *CCNE1* copy number (CN) gain (CN 2–5); three (21%) had CN amplification (CN > 5); and four (29%) had no alterations (Fig. 3B and table S10) (*40*). It is likely that *CCNE1* amplification in the pretreatment samples was acquired after prior treatment regimens, as *BRCA* mutation and *CCNE1* amplification are generally mutually exclusive (*2*). All four patients with clinical benefit also harbored *CCNE1* copy number gains/amplification and overexpression of its mRNA relative to those without clinical benefit (Fig. 3B and 5C). This finding suggests that tumors with a higher degree of replication stress may demonstrate a more durable response to CHK1i, regardless of *BRCA* mutation status or PARPi resistance. We also examined whether cyclin E1 protein levels may predict sensitivity to CHK1i. Notably, CHK1i sensitivity was not restricted to cells with cyclin E1 protein overexpression (fig. S6C), which is consistent with a previous report of prexasertib in HGSC cell lines (*18*). Additional studies have also suggested that CHK1i sensitivity is not dependent on only cyclin E1 in a subset of cancer cells, including HGSC cells (*41, 42*). These data further support our finding that increased levels of both *BLM* and *CCNE1*, rather than each independently, may be a better predictor of high replication stress and concomitant sensitivity to DNA repair inhibitors (fig. S5C).

### Tumor immune microenvironment shows a positive trend in patients with clinical benefit

Lastly, we investigated whether a correlation exists between the response of CHK1i and the prevalence of the immunoreactive (IMR) subtype associated with increased immune response, enhanced cytokine expression, and tumor-infiltrating lymphocytes (TILs) (*43-45*) in this *BRCA*-mutated HGSC cohort, as we previously reported tumor immune microenvironment (TIM) might have a potential predictive role in *BRCA*-wildtype HGSC for CHK1i sensitivity (*44*). Approximately half (8/15) of all evaluable patients showed profiles similar to IMR (fig. S8A), with high activation observed in one patient with CR, though this was not associated with PFS or clinical benefit (fig. S8B). Besides IMR, mesenchymal (MES) and proliferative (PRO) subtypes, characterized by increased stromal components and proliferation markers, respectively (*43*), were enriched in those without durable clinical benefit, in line with studies suggesting poorer prognosis associated with these subtypes (*2, 43*). Orthogonally, immunohistochemical analysis of CD3+ T lymphocyte and CD8+ cytotoxic T cell populations on tissue biopsies of patients with clinical benefit versus those with no clinical benefit show no significant difference (fig. S9A-B and table S6, suggesting that T-cell mediated response in the tumor microenvironment is an unlikely predictive biomarker for determining benefit to CHK1i in the *BRCA*-mutant setting.

## DISCUSSION

There is an urgent unmet need for novel therapeutic agents and relevant biomarkers to predict response in PARPi-resistant HGSC. In this study, high-throughput drug screens of both *BRCA*-mutant and *BRCA*-restored PARPi-resistant HGSC cell lines revealed high scores for agents that target replication fork stability and DNA replication, with the CHK1i prexasertib ranked high on the list. Prexasertib has shown clinical benefit previously in the treatment of recurrent *BRCA*-wildtype HGSC patients (*24*), as well as in preclinical models of PARPi-resistant HGSC (*18*). Here, we also observed considerable antitumor activity of CHK1i in PARPi-resistant HGSC cell lines irrespective of *BRCA* functional status. While our study demonstrated durable single agent activity of CHK1i in a proportion of this cohort of PARPi-pretreated HGSC patients with *BRCA* mutation, the results suggest that a biomarker approach that is able to improve the ORR in response to CHK1i is desperately needed.

Currently, there is no clearly defined replication stress biomarker to predict the response of CHK1i in the *BRCA*-mutant setting although *CCNE1* overexpression or copy number gain has been studied as a potential biomarker in HGSC for CHK1i sensitivity (*6, 24*). Also, the EVOLVE study (*39*) and others (*46, 47*) have found that *CCNE1* amplification or overexpression induces chemoresistance in ovarian cancer. But recent clinical studies have shown that *CCNE1* overexpression or copy number gain alone might not predict the sensitivity to cell cycle checkpoint blockade (*30, 48*). Additionally, a recent study by Konstantinopoulos *et al*. identified replication stress signatures for the ATRi berzosertib and gemcitabine sensitivity in HGSC patients (*29*). In this study, HGSC patients with high replication stress tumors benefited from gemcitabine alone, whereas those with low replication stress tumors benefited from the addition of berzosertib to gemcitabine (*29*). However, in our cohort, the replication stress signatures including *RB1* two-copy loss, *CDKN2A* two-copy loss, *KRAS* amplification, *ERBB2* amplification, *MYC* amplification, *NF1* mutations, and *MYCL1* amplification (*29*) were not associated with clinical benefit to CHK1i (fig. S7 and table S10), requiring further investigation.

In the current study, we found that mRNA overexpression of a RecQ helicase BLM along with replication stress marker *CCNE1* was markedly associated with increased clinical benefit to CHK1i in HGSC patients with *BRCA* mutation. Mechanistically, BLM is one of the key responders of stalled replication forks during replication stress (*49*). Preclinical studies have identified upregulation of BLM or BLM-positive ultrafine bridges in *CCNE1*-overexpressing primary human fallopian tube epithelial cells (*50*) and *RB1*-deficient osteoblastoma cells (*51*). Similarly, increased recruitment of BLM on stalled replication forks was observed in cells with high levels of RPA and single-stranded DNA (*52*), suggesting BLM’s critical function in replication fork stability, particularly under high levels of replication stress. Also, *BLM* copy number gain or increased *BLM* mRNA levels was shown in platinum-sensitive triple-negative breast cancer patients, suggesting its potential use in the clinic (*35*). Together, these reports along with our findings indicate the potential of BLM as a predictive biomarker for drugs targeting replication fork stabilization and our finding is the first demonstration of a replication fork-related biomarker in HGSC for CHK1i sensitivity.

Although several PARPi resistance mechanisms have been described in preclinical models, clinical data are relatively scarce and mostly confirm the prevalence of reversion mutations as a primary driver of PARPi resistance (*4, 25, 53*). As such, it is noteworthy that BRCA restoration had little correlation with the sensitivity to prior PARPi or platinum-based therapy and did not exclude a benefit from CHK1i in our cohort. A transcriptomic signature of replication stress has been reported as a predictive biomarker for ATR and WEE1 inhibitors in patient-derived pancreatic cancer cells and xenografts, in which response, similar to our findings, was not associated with HR deficiency (*54*). Thus, the use of replication stress and replication fork biomarkers might be more edifying toward predicting CHK1i and other cell cycle checkpoint inhibitors’ (*e*.*g*., ATR and WEE1 inhibitors) response when *BRCA* reversion mutations are factored in. Another factor that might be considered in this scenario is the immune milieu in the tumor microenvironment that has been previously shown by us and others (*44, 55, 56*) to be associated with clinical benefit to prexasertib in *BRCA*-wildtype HGSC. However, we did not observe the same association in the *BRCA*-mutant setting, further highlighting the complex molecular dynamics of PARPi resistance and CHK1i sensitivity.

Limitations of our study include the small size of our patient group and the progressive nature of PARPi resistance development. Also, all biomarker analyses are exploratory and hypothesis-generating in nature and warrant validation in a larger, prospective setting. Though cyclin E1 overexpression by immunohistochemistry staining has been associated with a higher ORR in platinum-resistant HGSC to the WEE1 inhibitor adavosertib (*57*), we were unable to examine whether protein overexpression of BLM or cyclin E1 in clinical samples, in addition to mRNA upregulation, correlates with clinical benefit to CHK1i because of the paucity of fresh core biopsy samples. In addition, there was only one patient who was PARPi-naïve, limiting the comparisons that could be made among PARPi-resistant and PARPi-sensitive subgroups in this cohort. Furthermore, the results of the present study in comparison with other clinical trials must be interpreted with caution given the lack of a standardized definition of clinical PARPi resistance, which we defined as progression on a prior PARPi at any time in the treatment life cycle after having an initial response to PARPi (*i*.*e*., acquired PARPi resistance). One way to address these challenges would be by increasing the number of clinical trials in the PARPi-resistant patient population with mandatory biopsies on enrollment along with collection of archival tissues, as it will be key to evaluate a dynamic measure of tumor replication stress status to provide the best treatment options. We contend that patients selected for high levels of replication stress along with replication fork related biomarkers should be considered for CHK1i-based therapy.

In summary, we demonstrate the therapeutic potential of CHK1i in a subgroup of *BRCA*-mutant HGSC, both in the preclinical as well as clinical settings, and report tumors with high dependence on replication fork stabilization may aid in identifying patients for CHK1i irrespective of PARPi resistance, warranting further prospective validation. Ultimately, our comprehensive molecular assessment of pretreatment tumors will help to guide the development of more personalized treatment regimens and improved patient stratification in HGSC with *BRCA* mutation.

## MATERIALS AND METHODS

### Preclinical Experiments

#### Cell lines

UWB1.289 (*BRCA1* mutation 2594delC) cell line was purchased from American Type Culture Collection (ATCC; #CRL-2945, Manassas, Virginia, USA). PEO1 (*BRCA2* mutation 5193C>G; #10032308-1VL) and PEO4 (*BRCA2* reversion mutation 5193C>T; #10032309-1VL) cell lines were purchased from Sigma-Aldrich (Saint Louis, Missouri, USA). The PARPi-resistant PEO1/OlaJR cell line was developed from parental PEO1 by culturing cells with olaparib from 5 μM to 40 μM over 3 to 4 months (*13*). Similarly, PARPi-resistant UWB/OlaR cells were developed from parental UWB1.289 by exposing them to olaparib from 0.5 μM to 20 μM over 12 months. Another PARPi-resistant PEO1/OlaR cell line is a gift from Dr. Bitler’s lab (*16*). PEO1 and UWB1.289 cells were grown in RPMI1640 medium with (+) L-glutamine supplemented with 10% fetal bovine serum, 0.01 mg/mL insulin, and 1% penicillin/streptomycin. PARPi-resistant cell lines were routinely maintained at 5 μM (PEO1/OlaR) or 20 μM (UWB/OlaR and PEO1/OlaJR) of olaparib. Cells were cultured without olaparib for at least 3 days before being used for the following experiments.

#### Chemical preparation

For *in vitro* assays, olaparib (#S1060), prexasertib (#S7178), and SRA737 (CCT245737, #S8253) were purchased from Selleck Chemicals (Houston, Texas, USA). 100 *μ*M of olaparib and 10 *μ*M of prexasertib and SRA737 were prepared as stocks in dimethyl sulfoxide (DMSO; #S-002-M, Sigma-Aldrich). All drugs were stored in aliquots at -80°C until use. For *in vivo* studies, prexasertib mesylate hydrate (prexasertib; #1234015-57-6, Invivochem LLc, Libertyville, Illinois, USA) was prepared in 20% CAPTISOL® (CyDex Pharmaceuticals, Inc., Lenexa, Kansas, USA), while olaparib (#V300, Invivochem LLc) was formulated in PBS containing 10% DMSO and 10% (w/v) 2-hydroxy-propyl-β-cyclodextrin as described previously (*18, 58*).

#### High-throughput single agent drug screening

High-throughput single agent drug screening was performed as previously reported (*15*). Briefly, 20 nl of MIPE 5.0 compounds were acoustically dispensed by Echo Acoustic Liquid Handler (Beckman Coulter Life Sciences, Indianapolis, Indiana, USA) into 1,536-well white tissue culture-treated plates. Each compound was plated at an 11-point concentration range with 1:3 dilution. Bortezomib, a proteasome inhibitor (final concentration 20 μM), was used as a positive control for cell cytotoxicity. PEO1, PEO4, and PEO1/OlaR were trypsinized and dispensed in 5 μL of growth medium using a Multidrop Combi dispenser at a density of 600 cells/well to allow for compounds to be present during exponential growth phase. Plates were incubated for 96 hours at standard incubator conditions and covered by a stainless steel gasketed lid to prevent evaporation. 3 μL of CellTiter-Glo® (Promega, Madison, Wisconsin, USA) were added to each well, and plates were incubated at room temperature for 15 minutes with a stainless-steel lid in place. Luminescence readings were taken using PHERAstar® (BMG Labtech, Ortenberg, Germany). Compound dose response curves were normalized to DMSO and empty well controls on each plate. The average Z-AUC was calculated to determine inactive and active drug responses. Drugs with average Z-AUC values less than -1.0 in PARPi-sensitive and -resistant HGSC cell lines in the MIPE 5.0 dataset were defined as ‘hits’ (*15*), indicating active drugs.

#### Cell proliferation assays: XTT and colony-forming assays

Cell survival ability was evaluated for the drug response of prexasertib and olaparib. For short-term cell survival, XTT assay (#X6493, Thermo Fisher Scientific, Pittsburgh, Pennsylvania) was performed. 2,000 cells/well were seeded in 96-well plates and treated with clinically attainable concentrations of prexasertib (0.125 nM to 100 nM (*17*)) and olaparib (2.5 to 50 *μ*M (*59*)) for 72 hours. The plates were measured by Synergy™ HTX Multi-Mode Microplate Reader with Gen5™ software (BioTek Instruments, Winooski, Vermont, USA). IC50 values were calculated using GraphPad Prism v7.1 (GraphPad Software, Inc., La Jolla, California, USA). The CI values were evaluated by CompuSyn software (ComboSyn, Inc., Paramus, New Jersey, USA). CI values less than 1 indicate synergism whereas greater than 1 indicate antagonism (*60*).

For long-term cell survival with prexasertib and/or olaparib treatment, colony-forming assay was used. 5,000 cells were seeded in 24-well plates and treated with prexasertib (0.5 nM for UWB1.289 and 5 nM for all other cell lines) and/or olaparib (10 *μ*M). Media and drugs were changed every three days for 12–15 days. Fixed colonies were stained with 0.01% (w/v) crystal violet in PBS. Colony images were scanned, and quantification of colony area percentage was done using Image J (NIH, Bethesda, Maryland, USA).

#### Immunoblotting

Active CHK1 (pCHK1-S296) and total CHK1 were measured to evaluate the target effects of CHK1i. Cells were treated with prexasertib (0.5 nM for UWB1.289 and 5 nM for all other cell lines). After 48-hour treatment, cells were collected for protein extraction and subjected to immunoblotting. Blots were visualized using the LI-COR Odyssey® Imaging System. BLM (#2742), CHK1 (#2360), pCHK1-S296 (#2349), ECL goat anti-mouse IgG HRP (#7076), and ECL goat anti-rabbit IgG HRP (#7074) antibodies were purchased from Cell Signaling Technology (Danvers, Massachusetts, USA). Cyclin E1 antibody (#ab74276) was purchased from Abcam (Waltham, Massachusetts, USA).

#### DNA fiber assay

DNA fiber assay was performed as described (*58*). Cells were labeled with 60 *μ*M CIdU (#I7125, Sigma-Aldrich) for 20 minutes, washed quickly and exposed to 500 *μ*M ldU (C6891, Sigma-Aldrich) for 10 minutes, and then treated with or without prexasertib (20 nM) and/or olaparib (20 *μ*M) for another 2 hours. Cells were collected and lysed with lysis buffer (1% sodium dodecyl sulfate, 100 mM Tris-HCl [pH 7.4], 50 mM EDTA). Labeled DNAs with CldU and IdU were stained with mouse anti-IdU primary antibody (1:250; #NBP2-44056, Novus Biological, Centennial, Colorado, USA) and rat anti-CldU primary (1:200; #NB500-169, Novus Biological), respectively. Anti-rat Alexa Fluor 488 (1:250; #A-11006, Thermo Fisher Scientific) and anti-mouse Alexa 594 (1:250; #A-11005, Thermo Fisher Scientific) were used for secondary antibodies. Images were captured with a Zeiss LSM 780 confocal microscope. Fiber length was measured using ImageJ software (NIH, Bethesda, Maryland, USA).

#### Assays for detecting DNA damage and HR repair status: alkaline comet assay and immunofluorescence staining of γH2AX and RAD51 foci

For DNA damage endpoints, DNA fragmentations and immunofluorescence staining of γH2AX foci formation were studied. Cells were treated with prexasertib (0.5 nM for UWB1.289 and 5 nM for all other cell lines) and/or olaparib (10 *μ*M) for 48 hours. DNA fragmentations were analyzed by alkaline comet assay according to the manufacturer’s instruction (Trevigen, Gaithersburg, Maryland, USA). The mean tail moment from three independent experiments (each experiment scored at least 100 cells/treatment) was calculated as an index of DNA damage by using CometScore Pro (TriTek Corporation, Sumerduck, Virginia, USA). For γH2AX foci formation, the cells were grown on Falcon™ chambered cell culture slides (Corning Inc., Oneonta, New York, USA). After drug treatment for 48 hours, cells were fixed in 4% paraformaldehyde (PFA) for 10 minutes, permeabilized with 0.25% Triton-X 100, and blocked with 1% bovine serum albumin in PBS for another 10 minutes. For RAD51 foci formation, cells were fixed in 3.7% PFA containing 2% sucrose and 0.5% Triton-X on ice as described before (*61*). All images were collected with an LSM 780 confocal microscope with a 63x/1.4 oil immersion objective. The number of γH2AX foci per nucleus was quantified by ImageJ. Cells with 5–15 γH2AX foci per nucleus and pan-nuclear γH2AX staining were counted (*20*). Cells with > 5 RAD51 foci per nucleus were counted as RAD51-positive cells (*61*).

#### BLM overexpression experiments

The GFP-BLM was a gift from Nathan Ellis (#80070, Addgene, Watertown, Massachusetts, USA) (*62*). Cells were seeded and transfected with GFP-BLM (100 ng) or GFP-empty vector in 96-well plates at a 3:1 FuGENE(r) HD Transfection Reagent: DNA ratio according to the manufacturer’s instruction (#E2311, Promega) for 48 hours. Cells were then treated with prexasertib for another 48 hours and subjected to cell proliferation assays.

#### Animal study

Xenograft murine models were used to evaluate the efficacy of prexasertib and olaparib *in vivo*. All animal procedures reported in this study that were performed by NCI-CCR affiliated staff were approved by the NCI Animal Care and Use Committee (ACUC) and in accordance with federal regulatory requirements and standards. All components of the intramural NIH ACU program are accredited by Association for Assessment and Accreditation of Laboratory Animal Care International (AAALAC). 5×10^6^ cells were suspended in 50 μl of cold PBS and mixed with 50 μl of Matrigel. A total of 100 *μ*l cell mixture was subcutaneously injected to 6-week-old female NSG mice (NCI, Frederick, Maryland, USA). The volume of tumor was measured once per week according to the formula V = ½ (length × width^2^). When tumors reached 50 mm^3^, mice were randomized into 3 groups (n = 5/group). Mice received vehicle or 8 mg/kg prexasertib by intraperitoneal (i.p.) injection twice daily (BID) for 3 days, followed by 4 days of no treatment for a total of 3 weeks (*58*). For combination studies, two study arms were added, and mice received 100 mg/kg olaparib orally once daily for 5 days, followed by 2 days of no treatment (*18*) with or without 8 mg/kg prexasertib i.p. BID for 3 days, followed by 4 days of no treatment for 3 weeks. All mice were sacrificed at the end of treatments.

#### Statistical analysis

Three independent biological replicates were performed in all experiments. Investigators were blinded during data collection and analysis. Data were analyzed using one-way ANOVA test and shown as mean ± SEM. The Pearson correlation coefficient was employed to analyze the correlation between basal BLM levels and colony-forming ability in cells with CHK1i treatment. All differences were considered statistically significant if p < 0.05. All statistical analyses were done using GraphPad Prism v7.1 (GraphPad Software).

### Clinical Trial

This open-label, single-center, single-arm phase II study was designed as a signal-seeking study with five independent patient cohorts: *BRCA*-mutant ovarian cancer; *BRCA*-wildtype HGSC (*24*); *BRCA*-wildtype triple negative breast cancer (*23*); *BRCA*-wildtype platinum-resistant HGSC; and *BRCA*-wildtype platinum-resistant HGSC without biopsiable disease. In this report, we present the final analysis of the *BRCA*-mutant ovarian cancer cohort with biomarker analyses. The full study methodology is described in the supplemental methods. Briefly, eligible patients received intravenous prexasertib monotherapy at 105 mg/m^2^ over one hour every 2 weeks in 4-week cycles (fig. S3A). Clinical response per RECIST v1.1 was assessed by the investigator every two cycles by computed tomography (CT) imaging or magnetic resonance imaging (MRI). Serum CA-125 response was investigated every cycle as a post-hoc exploratory end point and was defined as a 50% reduction during treatment with confirmation after 4 weeks according to GCIG criteria (*63*). Patients were evaluated for toxicity per Common Terminology Criteria for Adverse Events (CTCAE) v4.0. AEs were assessed before administration of the study drug on days 1 and 15 of cycle one, and before the start of every subsequent cycle. Transient (lasting ≤ 7 days) grade 3 or 4 neutropenia without fever or febrile neutropenia on cycle 1 without growth factor support did not require dose reduction or discontinuation of treatment. Patients received treatment until progression of disease, intercurrent illness, AEs not recovering to ≤ grade 1 within a 3-week period, or patient withdrawal of consent. Treatment interruptions of up to 7 days were permitted due to holidays, inclement weather preventing clinic attendance, toxic effects, or similar reasons.

The primary endpoint was investigator-assessed ORR (CR + PR) according to RECIST v1.1. Secondary endpoints included safety and toxicity evaluation per CTCAE v4.0; and PFS, defined as time from enrollment until the first documented disease progression according to RECIST v1.1 or death resulting from any cause.

The study was conducted using Simon’s optimal two-stage phase II trial design to rule out an unacceptably low 5% ORR in favor of an improved 25% ORR, with α=0.10 and β=0.10. These parameters were chosen to minimize the number of women exposed to a potentially inactive agent and to target a sufficiently high ORR to support moving into a definitive trial should this trial be positive. A response in 1 of the first 9 patients sufficed to move to the second stage of accrual, adding another 15 patients for a total of 24 patients. The regimen would be considered sufficiently interesting if ≥ 3/24 patients (12.5%) had a CR or PR, but the study was closed after enrolling 22 patients due to slow accrual. The null hypothesis of 5% was selected to accommodate the inclusion of heavily pretreated patients, based on the findings of the Gynecologic Oncology Group 0126 series of cancer trials, in which the proportion of patients with response was 3–4% (*64*). Under the null hypothesis, the probability of early termination was 63.0%. PFS was estimated using the Kaplan-Meier method beginning at the on-study date and continuing until progression or death without progression. Safety evaluation was based on all enrolled patients. Descriptive statistics were used to summarize the number and types of AEs. Patients considered non-evaluable had either no post-baseline CT scan or discontinued after less than 8 weeks without documented progression. Statistical tests for correlative studies utilized a two-sided significance level of 0.05. For gene expression profile analysis, we used the *limma* VOOM method followed by adjusting with Benjamini-Hochberg false discovery rate cut-off < 10% (adjusted p-value (q) < 0.1 indicates significance) for multiple hypothesis testing (*65*). This method allows for different levels of variability between genes and between samples and makes statistical conclusions more reliable (*65-67*). All patients provided written informed consent before enrollment. The study was conducted in accordance with ethical principles founded in the Declaration of Helsinki. The trial was approved by the Institutional Review Board of the Center for Cancer Research (CCR), NCI, USA (NCT02203513).

### Correlative Studies

For post-hoc correlative studies, we collected pretreatment CT- or ultrasound-guided fresh-frozen core biopsies (n = 15) and blood samples (n = 18) at baseline. Three patients had pretreatment biopsies cancelled due to safety reasons. Prespecified post-hoc exploratory objectives were investigation of potential predictive biomarkers of response to CHK1i in tumor and blood samples.

#### BRCA reversion mutation assay

cfDNA sequencing libraries were prepared from 5–10 ng of cfDNA input using the KAPA HyperPrep kit (Roche Sequencing and Life Science, Wilmington, Massachusetts, USA). Samples were individually barcoded using xGen UDI-UMI adapters (Integrated DNA Technologies, Coralville, Iowa, USA) and then pooled prior to multiplexed hybridization capture using xGen NGS Hybridization Capture for NGS target enrichment (Integrated DNA Technologies) targeting 200 kb of genomic territory derived from assorted coding segments of 68 cancer-associated genes. Samples were subsequently amplified, purified using AMPure beads (Beckman Coulter Life Sciences, Indianapolis, Indiana, USA), and quantified prior to sequencing on the Illumina NextSeq 500.

Samples were sequenced to no more than 50 million read pairs. Bioinformatics were performed by the University of Washington NGS Analytics Laboratory. Following demultiplexing and alignment, fgbio (Fulcrum Genomics, Boulder, Colorado, USA) was used to group reads by the unique molecular indices applied during adapter ligation prior to PCR. Families with fewer than five reads and those where greater than 5% of bases were identified as erroneous were discarded; the rest were collapsed into a single consensus sequence for each family, thereby reducing background noise due to sequencer error. In the post-processed sequence, bases where consensus could not be determined or where more than 10% of contributing reads did not support the consensus were masked to N. Collapsed families with more than 10% of their bases masked were also discarded. Samples were then realigned, and variants were called using VarDict (AstraZeneca-NGS, Cambridge, England). Alternate alleles that were supported by at least one read (with a base quality score > 10) and with a VAF > 0.001% were emitted to the initial VCF, which was filtered to a finalized list of mutations using custom filtration parameters. Large insertions and deletions were detected by manual inspection of regions adjacent to known germline alterations using Integrative Genomics Viewer (Broad Institute, Cambridge, Massachusetts, USA).

#### RNA sequencing (RNAseq) and whole exome sequencing (WES)

RNAseq and WES was performed using a HiSeq3000 sequencing system (Illumina, San Diego, California, USA) at the NCI CCR Sequencing Facility, Frederick National Laboratory for Cancer Research, as detailed previously (*44*). More specific details have been provided in supplemental methods.

## Data Availability

All data produced in the present work are contained in the manuscript.

## List of Supplementary Materials

Materials and Methods

figs. S1 to S10 for multiple supplementary figures

tables S3, S4, S6 for multiple supplementary tables

Data file Tables S1, S2, S5, S7, S8, S9, and S10 (Excel files)

References (63, 64, 68, 69)

## Acknowledgments

We thank Dr. Christina Annunziata, Ms. Mireya Gomez, Ms. Nicole Houston, Dr. Elise Kohn, Dr. Stanley Lipkowitz, Ms. Erin Villanueva, and Dr. Alexandra Zimmer for their contributions in clinic, Dr. Seth Steinberg for his assistance in statistical analysis of the clinical trial, and Dr. Nishanth Nair for his consultation on gene expression profile analysis. Eli Lilly supplied prexasertib to the NCI CCR under a cooperative research and development agreement.

## Author contributions

Conceptualization: NG, TH, JN, JL

Methodology: NG, TH, JN, JBR, CYC, KMW, DN

Investigation: NG, TH, JN, JL

Visualization: NG, TH, JN, DN, KMW, JBR

Funding acquisition: JL, EMS, NG, TH, KCC

Project administration: JL

Supervision: JL

Writing – original draft: NG, TH, JN, JL

Writing – review & editing: NG, TH, JN, DA, GZ, EJL, AM, ACM, EMS, MRR, TL, JBR, CYC, KMW, KCC, DN, JL

## Data and materials availability

All data are available in the main text or the supplementary materials.

