## Supplementary data for "BLM overexpression as a predictive biomarker for CHK1 inhibitor response in PARP inhibitor–resistant BRCA-mutant ovarian cancer"

**Supplementary Materials and Methods**
**Clinical Trial**
***Study design and participants***

Eligible patients were age ≥ 18 years and had histologically or cytologically confirmed recurrent high-grade serous ovarian cancer (HGSC), primary peritoneal cancer, and/or fallopian tube cancer with a documented deleterious germline or somatic *BRCA1* or *BRCA2* mutation. Patients must have had measurable disease by RECIST v1.1 and at least one lesion deemed safe for mandatory baseline percutaneous core biopsy. There were no restrictions on the number of prior treatment regimens, including platinum drugs and PARP inhibitors (PARPis). Other inclusion criteria were radiological progression after one or more lines of therapy, an ECOG performance status 0–2, and adequate organ and marrow function, defined as hemoglobin ≥ 10 mg/dL; leukocytes ≥ 3,000/mcL; absolute neutrophil count ≥ 1,500/mcL; platelet count ≥ 100,000/mcL; total bilirubin ≤ 1.5 × the upper limit of normal (ULN); alanine aminotransferase and aspartate aminotransferase ≤ 3 × ULN; and serum creatinine ≤ 1.5 × ULN or measured glomerular filtration rate ≥ 45 mL/min/1.73 $m^2$. Study exclusion criteria included concurrent anticancer therapy or any investigational anticancer therapy ≤ 4 weeks before the first dose of prexasertib (LY2606368); prior prexasertib or other cell cycle checkpoint kinase inhibitors; central nervous system metastases diagnosed within 1 year of enrollment; serious cardiac conditions; $QT_c$ interval > 470 msec on screening electrocardiogram; and uncontrolled intercurrent illnesses ≤ 28 days prior to start of the study. All patients provided written informed consent before enrollment. The study was conducted in accordance with ethical principles founded in the Declaration of Helsinki. The trial was approved by the Institutional Review Board of the Center for Cancer Research (CCR), National Cancer Institute (NCI), USA (NCT02203513).

***Procedures***

Eligible patients received intravenous prexasertib monotherapy at 105 $mg/m^2$ over one hour every 2 weeks in 4-week cycles (Supplementary Fig. 3a). Blood counts were repeated on day 8 of cycle 1 to establish the lowest absolute neutrophil count. Growth factor support was prohibited during the first dose. Laboratory assessments (including hematology, fasting serum chemistry, and urinalysis) and electrocardiogram were done within 24 hours before each study drug administration during cycle 1 and before the start of each subsequent 4-week cycle. Clinical response per RECIST v1.1 was assessed by the investigator every two cycles by computed tomography (CT) imaging or magnetic resonance imaging (MRI). Serum CA-125 response was investigated every cycle as a post-hoc exploratory end point and was defined as a 50% reduction during treatment with confirmation after 4 weeks according to GCIG criteria (*63*).

Patients were evaluated for toxicity per Common Terminology Criteria for Adverse Events (CTCAE) v4.0. AEs were assessed before administration of the study drug on days 1 and 15 of cycle one, and before the start of every subsequent cycle. Transient (lasting ≤ 7 days) grade 3 or 4 neutropenia without fever did not require dose reduction or discontinuation of treatment. Unresolved febrile neutropenia with growth factor support > 7 days, grade 3 or 4 anemia refractory to transfusion and growth factor support within 3 weeks, and grade 3 or 4 thrombocytopenia > 7 days or any thrombocytopenia requiring platelet transfusion for bleeding resulted in dose reduction to 80 $mg/m^2$ every two weeks for the remainder of the study. Patients received treatment until progression of disease, intercurrent illness, AEs not recovering to ≤ grade 1 within a 3-week period, or patient withdrawal of consent. Treatment interruptions of up to 7 days were permitted due to holidays, inclement weather preventing clinic attendance, toxic effects, or similar reasons. For safety reasons, prexasertib was not administered within 14 days of the previous dose.

### *Statistical analysis*

The study was conducted using Simon's optimal two-stage phase II trial design to rule out an unacceptably low 5% objective response rate (ORR) in favor of an improved 25% ORR, with $\alpha=0.10$ and $\beta=0.10$. These parameters were chosen to minimize the number of women exposed to a potentially inactive agent and to target a sufficiently high ORR to support moving into a definitive trial should this trial be positive. A response in 1 of the first 9 patients sufficed to move to the second stage of accrual, adding another 15 patients for a total of 24 patients. The regimen would be considered sufficiently interesting if ≥ 3/24 patients (12.5%) had a complete response (CR) or partial response (PR), but the study was closed early after enrolling 22 patients due to slow accrual and COVID19-related travel issues. The null hypothesis of 5% was selected to accommodate the inclusion of heavily pretreated patients, based on the findings of the Gynecologic Oncology Group 0126 series of cancer trials, in which the proportion of patients with response was 3–4% (*64*). Under the null hypothesis, the probability of early termination was 63%. PFS was estimated using the Kaplan-Meier method beginning at the on-study date and continuing until progression or death without progression. Safety evaluation was based on all enrolled patients. Descriptive statistics were used to summarize the number and types of AEs. Patients considered non-evaluable had either no post-baseline CT scan or discontinued after less than 8 weeks without documented progression. All statistical tests for correlative studies utilized a two-sided significance level of 0.05 and are reported without adjustment for multiple comparisons due to the small study cohort and exploratory nature of this analysis. This ongoing trial is registered with ClinicalTrials.gov (NCT02203513).

### *RNA sequencing (RNAseq)*

Total RNA was isolated from pretreatment fresh-frozen core biopsies using the RNeasy microkit (Qiagen, Hilden, Germany). RNA quality was evaluated using the Agilent Bioanalyzer 2100 and RNA integrity number (RIN) values were ensured to be > 8.0. For total RNAseq, each sample (20 to 100 ng) was preprocessed with NEBnext rDNA depletion kit (New England Biolabs, Ipswich, Massachusetts, USA) to remove ribosomal RNA, then barcoded and pooled to ensure at least 100 million reads per sample on a HiSeq3000 sequencing system (Illumina). The human reference genome hg38 and Gencode V30 were used to align reads, and gene expression data were generated as counts per million mapped reads (CPM) values. Quality check of sample and sequencing outputs were performed by the CCR sequencing facility and CCR Collaborative Bioinformatics Resource at NCI, Bethesda, Maryland, USA. $\text{Log}_2$ of CPM values were used throughout. Differential expression of genes (DEG) and Reactome, KEGG, and Hallmark pathway analyses were conducted using GSEA software (Broad Institute, Inc., Massachusetts Institute of Technology, and Regents of the University of California). Median and 95% CI for $\log_2$ of CPM gene expression across clinical benefit and no clinical benefit groups were calculated and a Wilcoxon test was used to determine significantly different gene expressions at a p-value of 0.05. HGSC subtype was determined using a validated single sample Gene Set Enrichment Analysis (ssGSEA) signature as has been previously described (*68*).

### *Immunohistochemistry (IHC) staining of CD3 and CD8*

Formalin-fixed and paraffin-embedded samples from pretreatment biopsies were available in 15 patients with RECIST response. Hematoxylin and eosin (H&E) staining of the unstained slides was performed and evaluated to confirm the pathologic diagnosis of HGSC. Automated CD3 and CD8 staining were performed on the Leica Bond RX (Leica Biosystems, Buffalo Grove, Illinois, USA). Slides were baked and dewaxed online followed either by low pH Sodium Citrate buffer antigen retrieval for 20 minutes at 100°C (for CD3), or by high pH EDTA buffer antigen retrieval for 20 minutes at 100°C (for CD8). Endogenous peroxidase was blocked, then either anti-CD3 clone EP204 LN10 (CD3-565-L-CE, Leica Biosystems) was applied for 30 minutes at a concentration of 12.4 µg/mL at room temperature (for CD3), or anti-CD8 clone 4B11 (CD8-4B11-L-CE, Leica Biosystems) was applied for 15 minutes at a concentration of 19.2 µg/mL at room temperature (for CD8). Detection was performed using the Bond Polymer Refine Kit (DS9800, Leica Biosystems). Slides were counterstained, dehydrated through a graded-alcohol, and coverslipped using Ecomount (5082832, Biocare Medical, Walnut Creek, California, USA).

***Slide scanning and digital image analysis***

IHC stained slides were scanned at 20x objective equivalent (0.49 microns/pixel) using the Hamamatsu NanoZoomer S360 whole slide scanner (Hamamatsu City, Japan). Image compression type was JPEG with a quality factor of 80. Each image was annotated for regions of tumor and non-tumor by the study pathologist (A.C-M.). Immune cell signals were detected in each region of non-necrotic tissue by digital analysis (cytonuclear IHC module in HALO v3.1, indica labs; Albuquerque, New Mexico, USA) and reported as cell density (positive cell number per $mm^2$ tissue area).

***Kaplan-Meier survival analysis***

The prognostic value of *BLM* and *CCNE1* mRNA expression was evaluated using an online database, Kaplan-Meier Plotter (www.kmplot.com) (*69*), which contained gene expression data and survival information of ovarian cancer patients. To analyze the progression-free survival (PFS) of patients with ovarian cancer, patient samples were split into two groups by median expression (high versus low expression) and assessed by a Kaplan-Meier survival plot, with the hazard ratio with 95% confidence intervals and logrank p value.

***Whole exome sequencing (WES)***

Library preparation was performed using Agilent SureSelect Human All Exon V7 target enrichment kit (Agilent Technologies, Santa Clara, California, USA). FASTQ files from sequencing were processed using the CCR Collaborative Bioinformatics Resource (CCBR) WES pipeline. Reads were trimmed using Trimmomatic v0.39 and mapped to the hg38 reference genome using BWA-MEM v0.7.17. The GATK4 Best Practices Workflow was followed for subsequent variant calling using GATK v4.2.0.0. Aligned BAM files were used to perform Base Quality Score Recalibration followed by somatic variant calling using Mutect2. A panel of normals was developed from normal The Cancer Genome Atlas (TCGA) samples; internal controls (patients' matching normal DNA from peripheral blood mononuclear cells) were used as reference for the Mutect2 step. Variants were annotated to COSMIC v92, ClinVar v2020-04-19, gnomAD v3.0, and ExAC v03 databases using ANNOVAR v2019-10-24. Copy number variants were called using a combination of Control-FREEC v11.6 and Sequenza v3.0.0. Sequenza was used as input into Control-FREEC to calculate tumor purity. Somatic variants called were included if they had a minimum of one read with at least 20x depth in the tumor sample and a minimum frequency of

0.05 in the tumor sample. Stringent filtering using population allele frequencies was applied to remove as many potential germline variants as possible, where mutations with a total allele frequency < 0.001 in the Genome Aggregation Database (gnomAD) and ExAC databases were removed. Somatic mutations were processed and visualized using the R/Bioconductor maftools package v2.8.05. The variants were visualized using oncoplots. The enrichment of variants was tested between the clinical benefit patients compared to patients without a clinical benefit using a Fisher's Exact test. Overall quality check of sample and sequencing outputs was performed by the CCR sequencing facility and CCBR at NCI, Bethesda, Maryland, USA.

**Supplementary figures and legends**

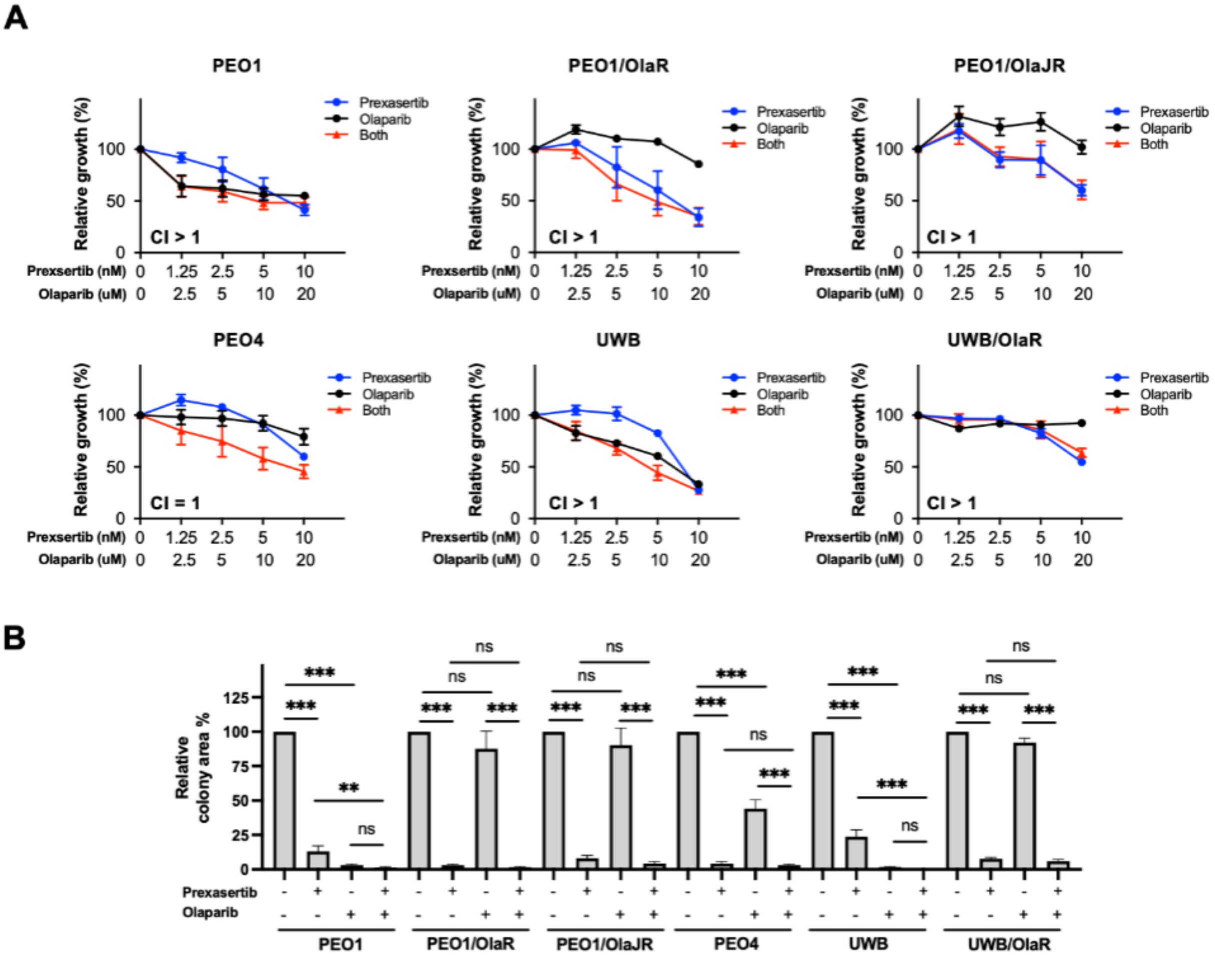


**Fig. S1. The addition of olaparib did not further increase CHK1i-induced cytotoxicity in PARPi-resistant *BRCA*-mutant HGSC cells (related to Fig. 1).** (**A**) Cell growth in multiple HGSC cell lines including parental (*BRCA*-mutant PEO1 and UWB), acquired PARPi-resistant (PEO1/OlaR, PEO1/OlaJR, and UWB/OlaR), and *de novo* PARPi-resistant (PEO4) cells were examined using XTT assay. Cells were treated with CHK1i prexasertib and/or PARPi olaparib at indicated doses for 72 hours. The CI quantitatively indicates additive/synergistic (CI < 1) or antagonism effect (CI ≥ 1). (**B**) Cells were seeded at low density and treated with prexasertib and/or olaparib grown for 12–15 days as described in Fig. 1C. The quantification was performed by ImageJ software. All data were repeated at least in triplicate and shown as mean ± SEM. *, $p < 0.05$; **, $p < 0.01$; ***, $p < 0.001$. Abbreviations: CHK1i, CHK1 inhibitor; CI, combination index; HGSC, high-grade serous ovarian cancer; PARPi, PARP inhibitor; UWB, UWB1.289.

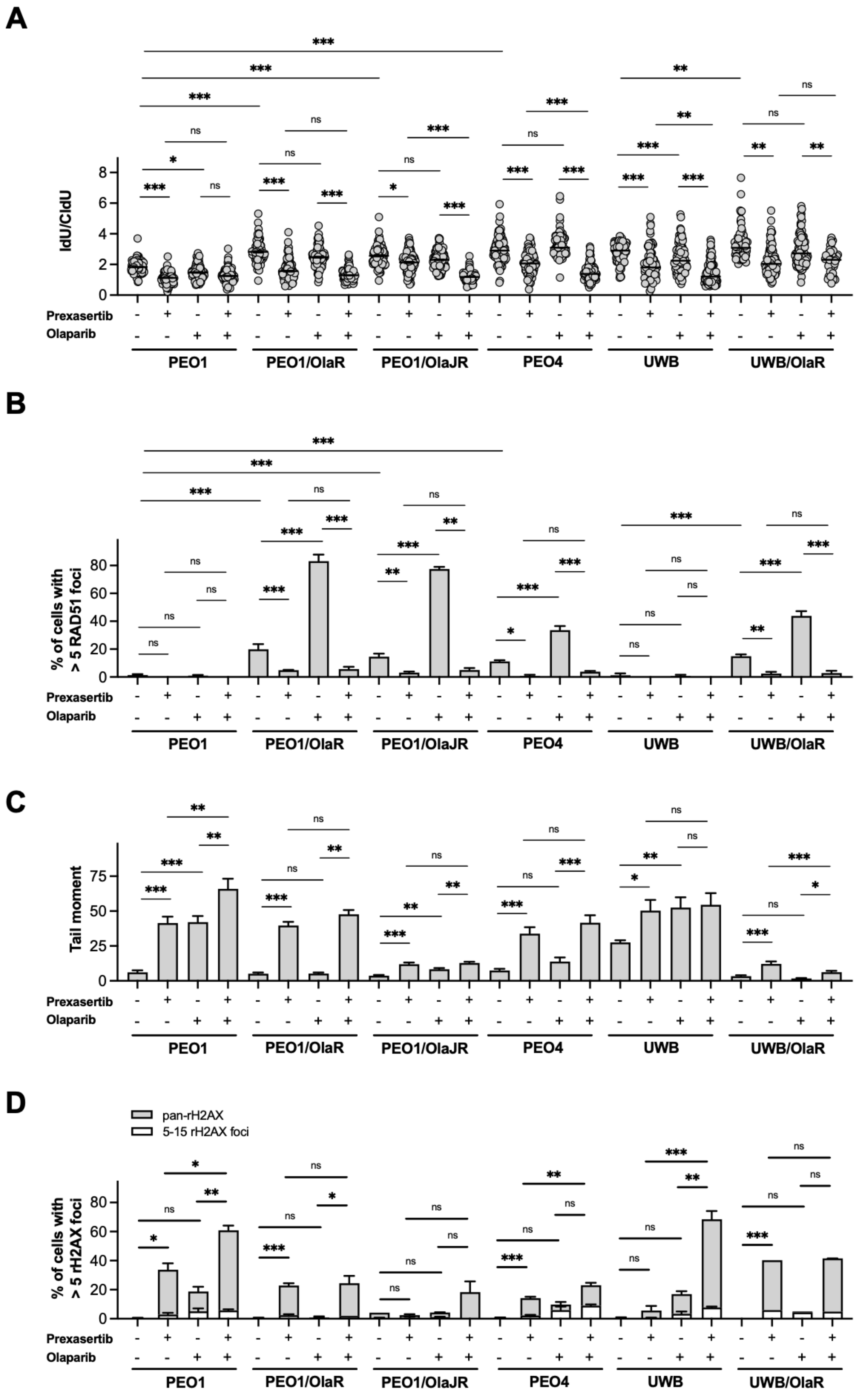
A
B
C
D
IdU/CldU
% of cells with > 5 RAD51 foci
Tail moment
% of cells with > 5 rH2AX foci
pan-rH2AX
5-15 rH2AX foci
Prexasertib
Olaparib
PEO1
PEO1/OlaR
PEO1/OlaJR
PEO4
UWB
UWB/OlaR

**Fig S2. The addition of olaparib does not further increase CHK1i-induced replication fork destabilization and DNA damage in PARPi-resistant *BRCA*-mutant HGSC cells (related to Fig. 2).** (**A**) DNA fiber assays were performed to measure replication fork stabilization in PARPi-naïve (*BRCA*-mutant PEO1 and UWB), *de novo* PARPi-resistant (PEO4), and acquired PARPi-resistant (PEO1/OlaR, PEO1/OlaJR, and UWB/OlaR) *BRCA*-mutant HGSC cell lines as described in Fig. 2a. Dot plots of IdU to CldU tract length ratios for individual replication forks in treated cells are shown. (**B**) HR restoration status was assessed by immunofluorescence imaging of RAD51 foci mentioned in Fig. 2B. Cells with > 5 RAD51 foci indicate RAD51-positive cells. (**C-D**) For DNA damage endpoints, alkaline comet assay (**C**) and immunofluorescence imaging of γH2AX foci (**D**) were performed as described in Fig. 2C-D. Cells were treated with prexasertib and/or olaparib for 48 hours. Mean tail moment is plotted (**C**). The percentage of cells with 5–15 γH2AX foci indicating DNA-double strand breaks and pan-γH2AX nuclear staining representing apoptosis are shown (**D**). All data were repeated at least in triplicate and shown as mean ± SEM. *, $p < 0.05$; **, $p < 0.01$; ***, $p < 0.001$; ns, not significant. Abbreviations: CHK1i, CHK1 inhibitor; CldU, 5-chloro-2′-deoxyuridine; HR, homologous recombination; IdU, 5-Iodo-2′-deoxyuridine.; PARPi, PARP inhibitor; UWB, UWB1.289.

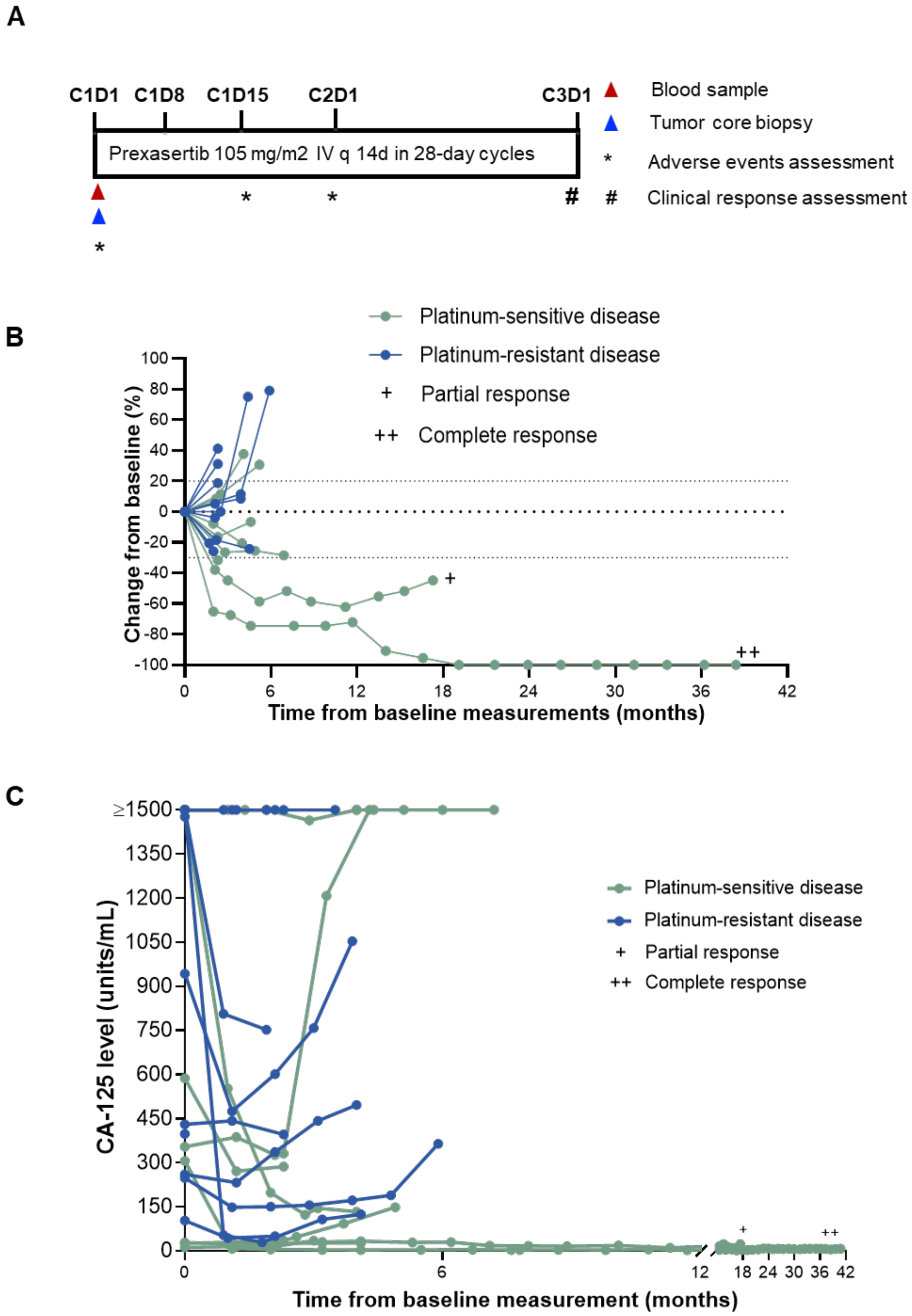


**Fig. S3. Clinical trial study schema, tumor measurements, and CA-125 levels.** (**A**) Study schema. (**B**) Baseline and serial tumor measurements in 18 evaluable patients. (**C**) Baseline and serial CA-125 measurements in 18 patients with at least one serum CA-125 level collected, 14 of whom were evaluable for CA-125 response based on GCIG criteria. In (**B**) and (**C**), the complete response patient is PARPi-naïve, whereas all other patients received prior PARPi. Abbreviations: C1D1, cycle 1 day 1; C1D8, cycle 1 day 8; C1D15; cycle 1 day 15; C2D1, cycle 2 day 1; C3D1, cycle 3 day 1; CA-125, cancer antigen 125; IV, intravenous.

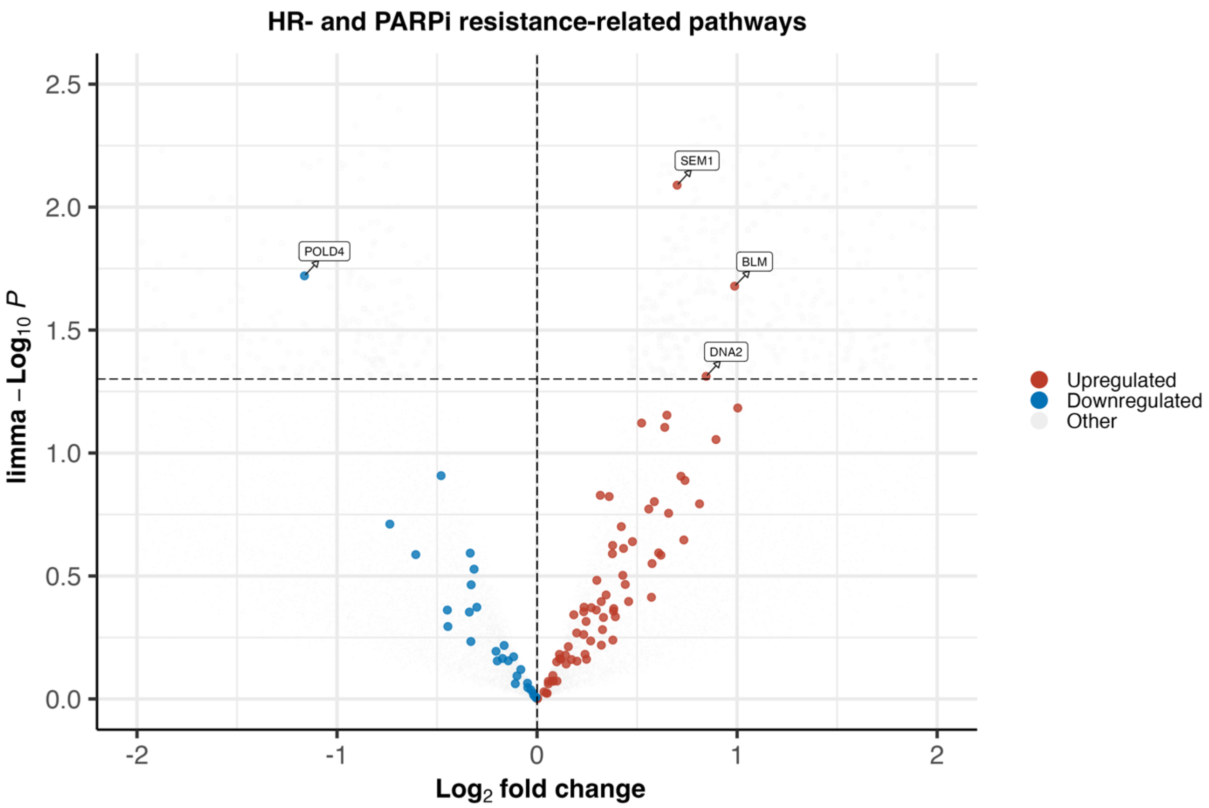


**Fig. S4. HR repair- and PARPi-resistance-related genes are largely not associated with clinical benefit to CHK1i.** Volcano plot showing gene set enrichments among pretreatment biopsy samples from patients with clinical benefit and no clinical benefit. The x-axis shows $\log_2$ fold change (>0 indicates genes enriched in patients with clinical benefit, while <0 indicates genes enriched in patients with no clinical benefit). The y-axis shows limma-$\log_{10}$ p values, and the horizontal dotted line represents a significance threshold of $p = 0.05$. Gene sets in the upper right and left quadrants are significantly enriched in the clinical benefit group and no clinical benefit group, respectively. Selected gene sets related to DNA repair are highlighted. Abbreviations: CHK1i, CHK1 inhibitor; HR, homologous recombination; PARPi, PARP inhibitor.

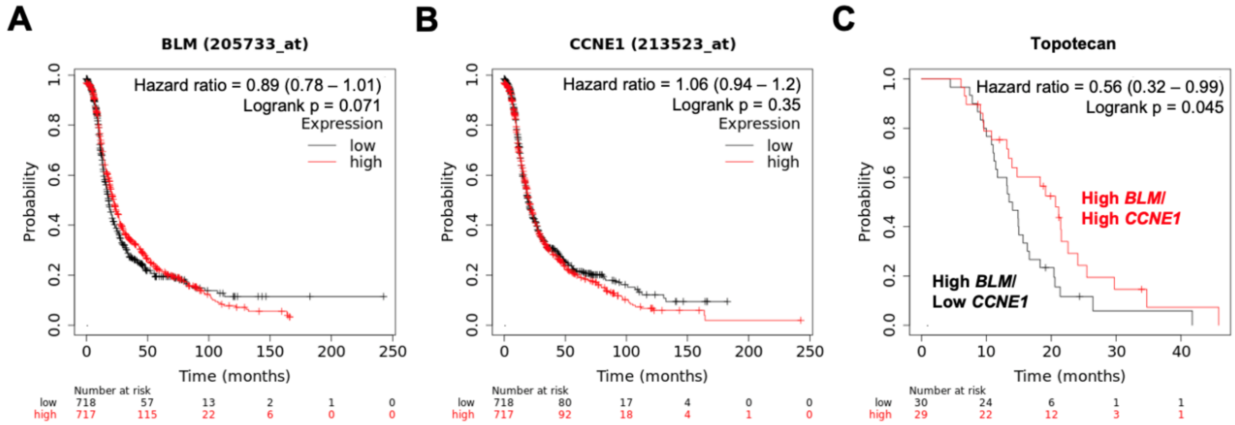


**Fig. S5. Effect of *BLM* and *CCNE1* expressions on survival rate of ovarian cancer patients.** (**A-B**) The expressions of either *BLM* (A) or *CCNE1* (B) by themselves do not significantly associate with PFS in ovarian cancer patients (n = 1,435). (**C**) High co-expression of *BLM* and *CCNE1* (n = 30) shows better PFS compared to those with low *CCNE1* and high *BLM* (n = 29) in patients treated with DNA replication inhibitor topotecan (Kmplot.com, accessed on June 1, 2022). Abbreviations: PFS, progression-free survival.

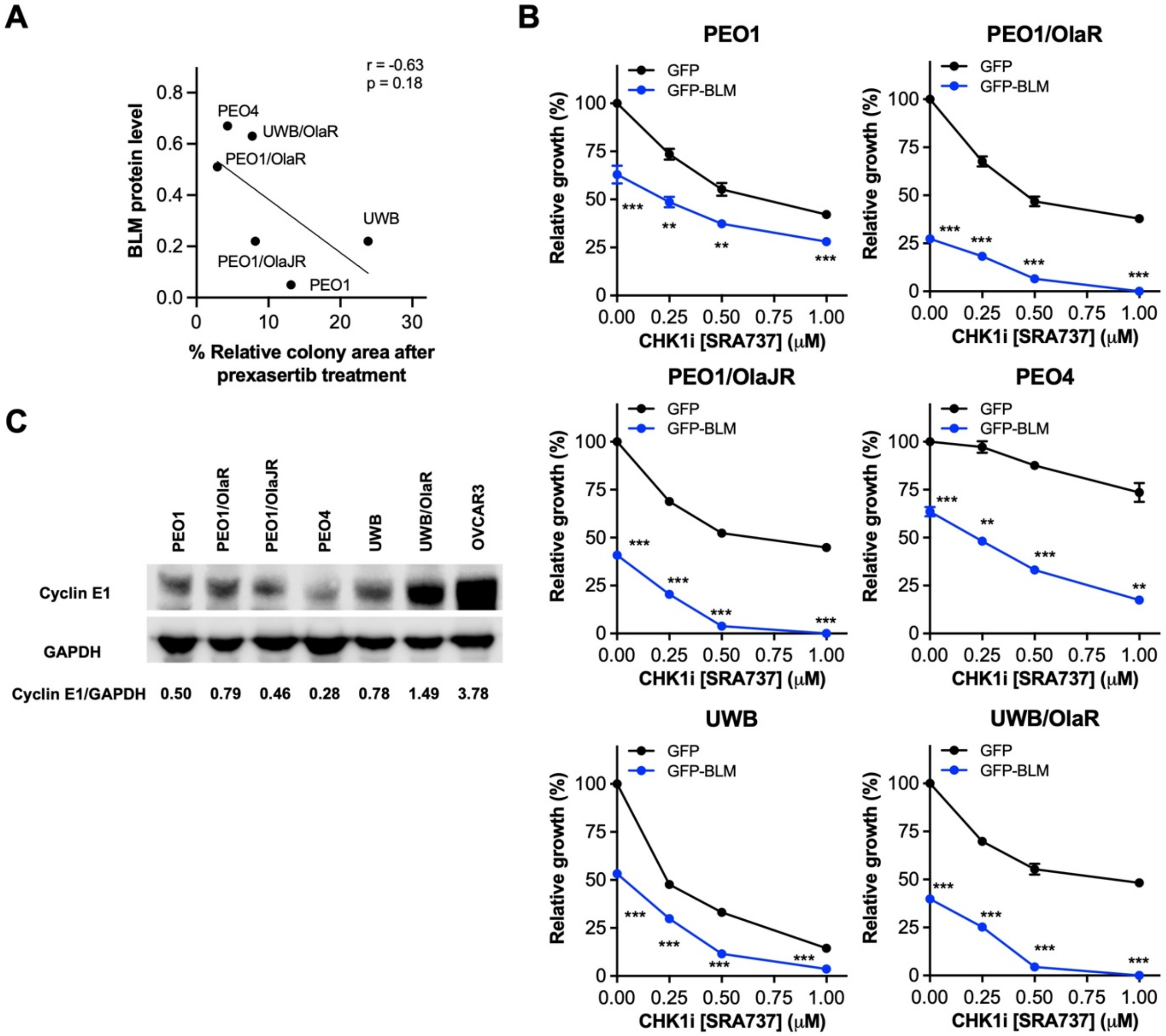


**Fig. S6. The baseline levels of BLM may be correlated with the sensitivity of CHK1i prexasertib in PARPi-sensitive and -resistant HGSC cells (related to Fig. 6). (A)** The Pearson correlation coefficient (r) between basal BLM levels (Fig. 6a) and colony-forming ability of cells with prexasertib treatment (Fig. 1d) is shown. (**B**) Cells transfected with BLM overexpression plasmids for 48 hours were treated with or without a specific CHK1i SRA737 for another 48 hours. Cell viability was measured by XTT assay. Experiments were repeated at least in triplicate and data are shown as mean ± SEM. **, $p < 0.01$; ***, $p < 0.001$. (**C**) Basal Cyclin E1 protein expression of each cell line is shown. *CCNE1*-amplified OVCAR3 was used as a positive control. Abbreviations: CHK1i, CHK1 inhibitor; PARPi, PARP inhibitor, UWB, UWB1.289.

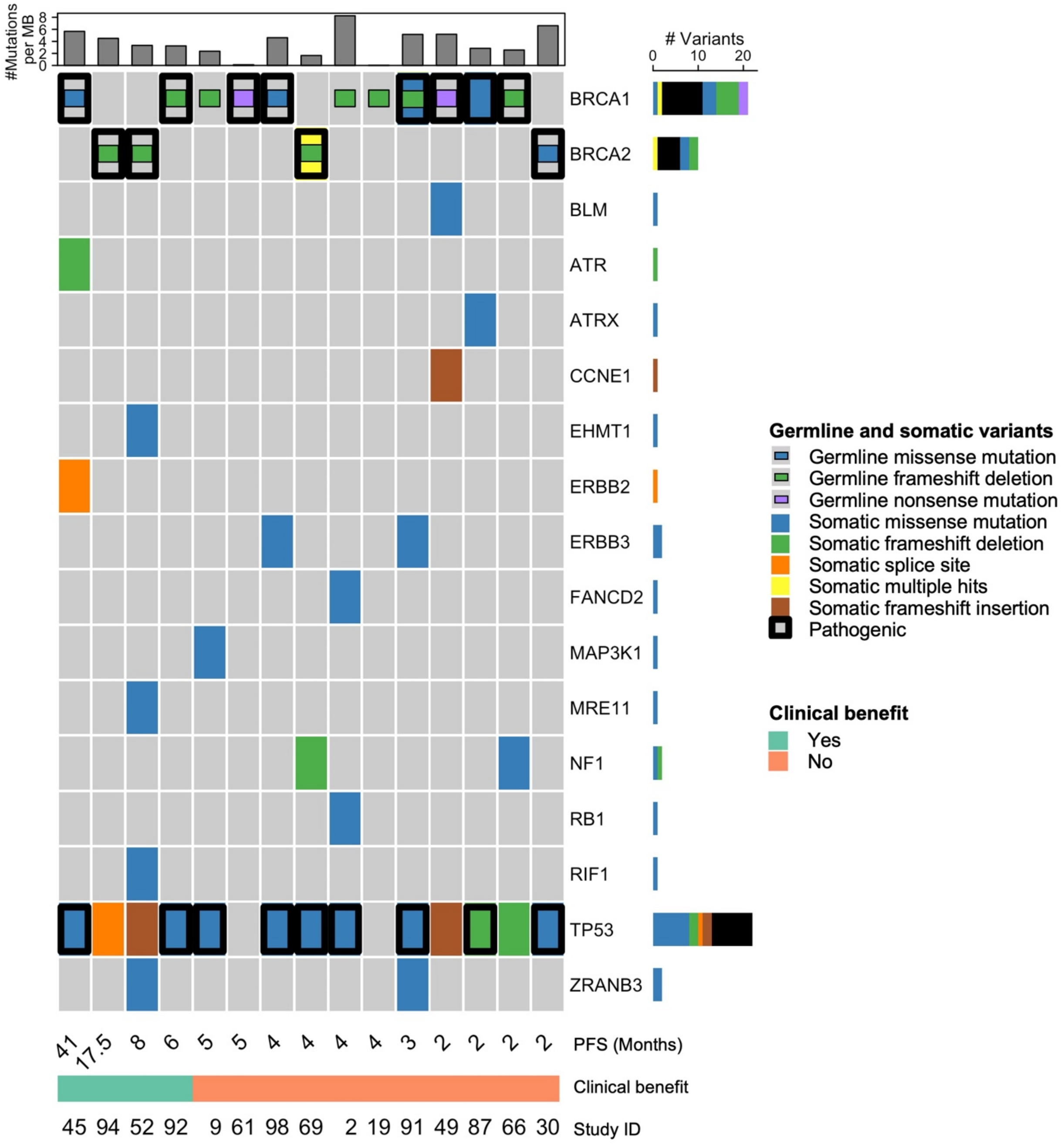


**Fig. S7. Germline and somatic variants in homologous recombination repair-related genes from whole exome sequencing.** Heatmap summarizing mutation burden, germline, and somatic variants derived from whole exome sequencing of patient tumors. Details of germline and somatic variants can be found in Table S5. We also confirmed the presence of germline/somatic *BRCA1* and *BRCA2* mutations in the available samples as reported in the CLIA-certified comprehensive commercial BRCA and/or HRD testing provided upon study enrollment (related to Table 1). In addition, we identified a somatic *BRCA1* mutation in one patient that was not reported in commercial testing, and we updated the *BRCA* mutation status in subsequent analyses (study ID 91; germline *BRCA1* mutation c.5329dup; somatic *BRCA1* mutation c.G140T). Study ID 45 is the complete responder and is PARPi-naïve, whereas all other patients received prior PARPi.

Abbreviations: CLIA, Clinical Laboratory Improvement Amendments; HRD, homologous recombination deficiency; PFS, progression-free survival.

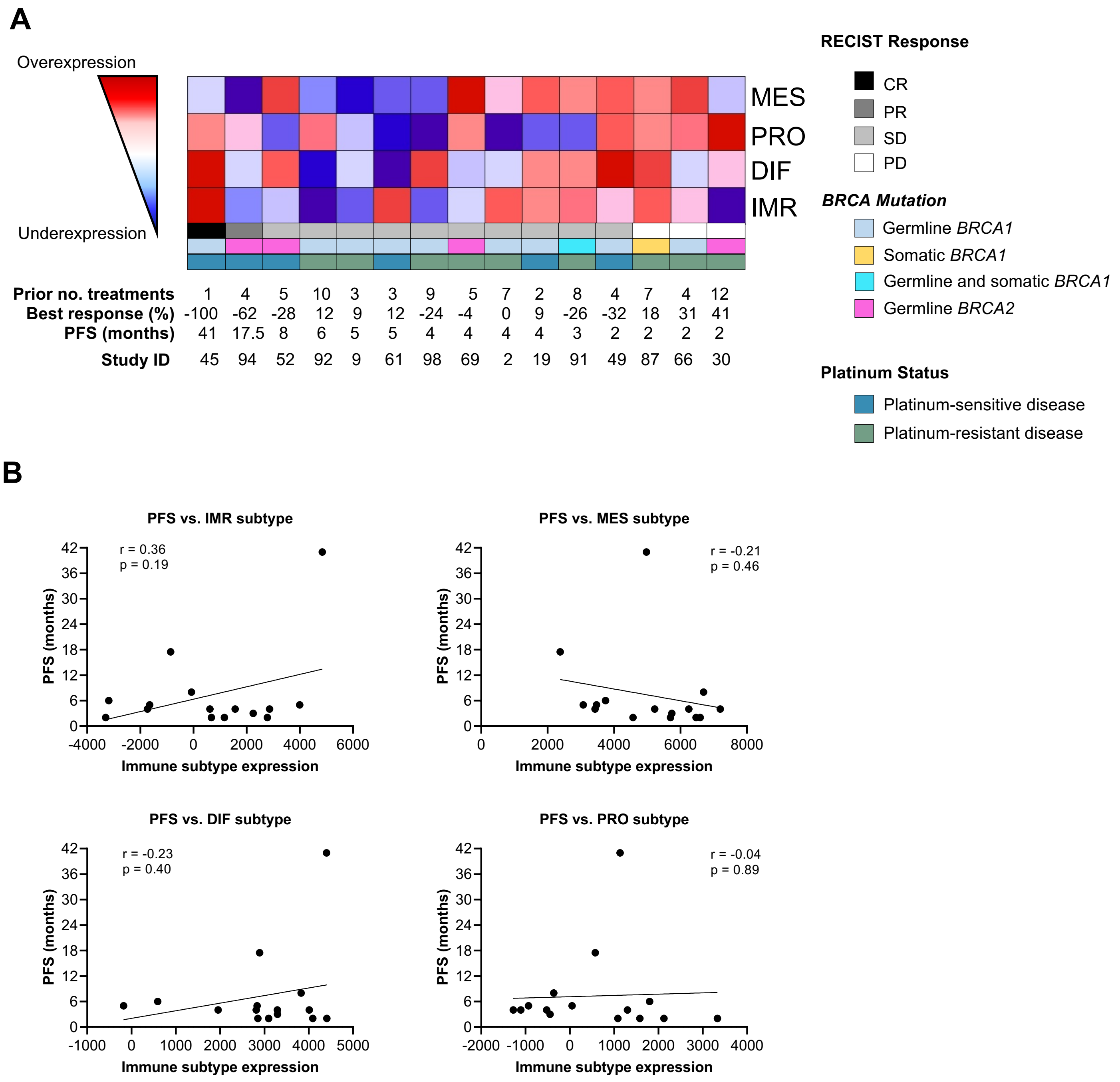


**Fig. S8. Immune expression subtypes are not significantly correlated with progression-free survival**. (**A**) Transcriptomic analysis of four HGSC molecular subtypes in 15 available pretreatment biopsies. Red indicates greater expression and blue indicates lower expression, with different shades of red and blue representing varying degrees of increased and decreased expression, respectively. Study ID 45 is PARPi-naïve, whereas all other patients received prior PARPi. (**B**) The mRNA expressions of different immune expression subtypes were done by

RNAseq. The correlation between PFS and each immune expression subtype was analyzed using GraphPad Prism v.7.1. Abbreviations: cfDNA, cell-free DNA; CR, complete response; DIF, differentiated; HGSC, high-grade serous ovarian cancer; IHC, immunohistochemistry; IMR, immunoreactive; MES, mesenchymal; PD, progression of disease; PFS, progression-free survival; PR, partial response; PRO, proliferative; SD, stable disease.

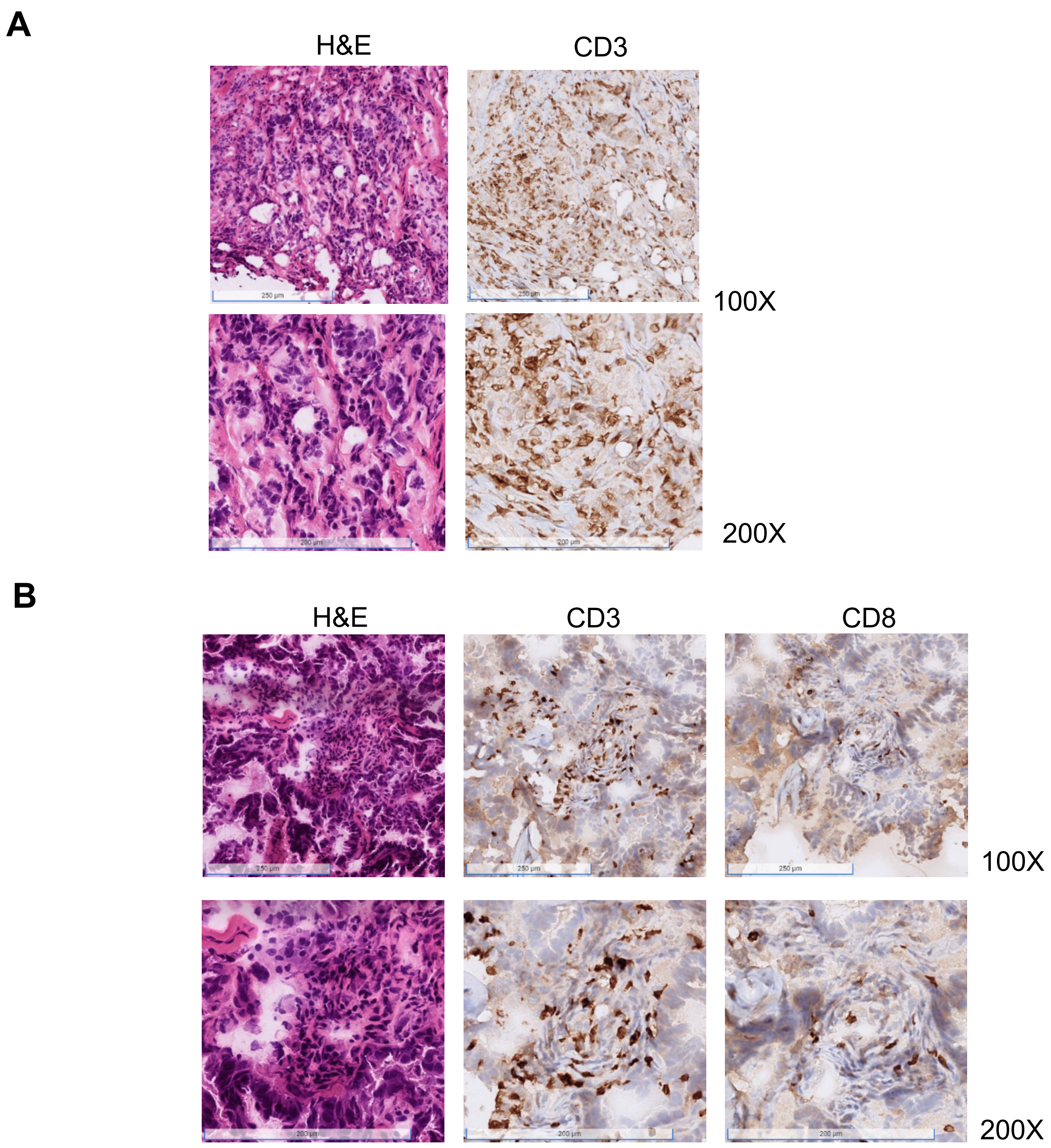


**Fig. S9. Representative images of immunohistochemical staining of CD3+ and CD8+ T lymphocytes.** (**A**) Immunohistochemical staining (H&E, CD3+ T lymphocytes) of pretreatment biopsies in the patient with CR. (**B**) Immunohistochemical staining (H&E, CD3+ T lymphocytes, and CD8+ cytotoxic T cells) of pretreatment biopsies in the patient with PR. Abbreviations: CR, complete response; H&E, hematoxylin and eosin; PR, partial response.

**Table S1. Whole compound list of drug screen in PEO1, PEO1/OlaR, and PEO4 cell lines (Please see Excel file).**

**Table S2. Oncology drug hits across PEO1, PEO1/OlaR, and PEO4 cell lines (Please see Excel file).**

**Table S3. All other treatment-related adverse events**

| Adverse event | Maximum grade in all patients (n = 22) | | |
|---|---|---|---|
| | 1–2 | 3 | 4 |
| **Non-hematologic** | **Number of patients (% total)** | | |
| **ALT/AST increase** | 4 (18%) | 0 | 0 |
| **Alkaline phosphatase increase** | 1 (5%) | 0 | 0 |
| **Alopecia** | 1 (5%) | 0 | 0 |
| **Arthralgia** | 1 (5%) | 0 | 0 |
| **Ascites** | 0 | 1 (5%) | 0 |
| **Back pain** | 2 (9%) | 0 | 0 |
| **Bone pain** | 1 (5%) | 0 | 0 |
| **Bruising** | 1 (5%) | 0 | 0 |
| **Chills** | 1 (5%) | 0 | 0 |
| **Cognitive disturbance** | 1 (5%) | 0 | 0 |
| **Creatinine increase** | 2 (9%) | 0 | 0 |
| **Cystitis noninfective** | 1 (5%) | 0 | 0 |
| **Dizziness** | 1 (5%) | 0 | 0 |
| **Dysgeusia** | 2 (9%) | 0 | 0 |
| **Fecal incontinence** | 1 (5%) | 0 | 0 |
| **Flu-like symptoms** | 1 (5%) | 0 | 0 |
| **Flushing** | 1 (5%) | 0 | 0 |
| **Gastritis** | 1 (5%) | 0 | 0 |
| **Hoarseness** | 1 (5%) | 0 | 0 |
| **Hyperhidrosis** | 1 (5%) | 0 | 0 |
| **Hypomagnesemia** | 1 (5%) | 0 | 0 |
| **Hyponatremia** | 3 (14%) | 0 | 0 |
| **Infusion-related reaction** | 1 (5%) | 0 | 0 |
| **Malaise** | 2 (9%) | 0 | 0 |
| **Myalgia** | 2 (9%) | 0 | 0 |
| **Palpitations** | 1 (5%) | 0 | 0 |
| **Sinus bradycardia** | 1 (5%) | 0 | 0 |
| **Sinusitis** | 1 (5%) | 0 | 0 |
| **Skin infection** | 2 (9%) | 0 | 0 |
| **Vaginal dryness** | 1 (5%) | 0 | 0 |
| **Weight loss** | 1 (5%) | 0 | 0 |

Patients could be counted under more than one preferred term.
Abbreviations: ALT, alanine transaminase; AST, aspartate aminotransferase.

**Table S4. *BRCA* reversion mutations in tissue and cfDNA**

| Patient Study ID | *BRCA* Mutation | Reversion Detected in Tissue | Reversion Detected in cfDNA | ctDNA Detected | Best RECIST Response | Clinical Benefit (CR + PR + SD ≥ 6 months) |
|---|---|---|---|---|---|---|
| 2 | BRCA1 c.68_69del | negative | negative | positive | SD | No |
| 3 | BRCA2 c.5946del | NA | positive | positive | SD | No |
| 9 | BRCA1 c.2188del | positive | positive | positive | SD | No |
| 19 | BRCA1 c.68_69del | negative | negative | positive | SD | No |
| 30 | BRCA2 c.9371A>T (p.N3124I) | negative | not evaluable | not detected | PD | No |
| 34 | BRCA1 c.5329dup | NA | negative | positive | PD | No |
| 45 | BRCA1 c.5186C>A (p.Ala1729Glu) | negative | negative | positive | CR | Yes |
| 49 | BRCA1 c.2603C>G (p.S868*) | negative | negative | positive | SD | No |
| 52 | BRCA2 c.4449del | positive | positive | positive | SD | Yes |
| 61 | BRCA1 c.3598C>T (p.Q1200*) | negative | positive | positive | SD | No |
| 62 | BRCA1 c.68_69del | NA | negative | positive | SD | No |
| 66 | BRCA1 c.798_799del | negative | negative | positive | PD | No |
| 69 | BRCA2 c.6275_6276del | positive | positive | positive | SD | No |
| 87 | BRCA1 c.140G>T (p.C47F) | negative | not evaluable | not detected | PD | No |
| 91 | BRCA1 c.5329dup | not evaluable | negative | positive | SD | No |
| 92 | BRCA1 c.1961del | positive | positive | positive | SD | Yes |
| 94 | BRCA2 c.8575delC | positive | not evaluable | not detected | PR | Yes |
| 98 | BRCA1 c.181T>G (p.C61G) | negative | negative | positive | SD | No |

Study ID 45 is PARPi-naïve, whereas all other patients received prior PARPi.

Abbreviations; CR, complete response; PD, progression of disease; PR, partial response; SD, stable disease.

**Table S5. Germline and somatic variants in homologous recombination repair-related genes from whole exome sequencing (Related to Supplementary Fig. 7, please see Excel file).**

**Table S6. Immunohistochemical analysis of CD3 and CD8 cell density**

| Best Response | Evaluable Patients * | CD3 cells/mm$^2$ (mean) | CD8 cells/mm$^2$ (mean) | Ratio CD8:CD3 cells/mm$^2$ (mean) |
|---|---|---|---|---|
| **PD** | 3 | 214 | 24 | 0.08 |
| **SD** | 5 | 846 | 87 | 0.1 |
| **PR** | 1 | 276 | 55 | 0.2 |
| **CR** | 1 | 839 | N/A | N/A |
| **Clinical Benefit** | | | | |
| **Yes** | 3 | 382 (n = 3) | 31 (n = 2) ** | 0.2 (n = 2) ** |
| **No** | 7 | 691 | 72 | 0.08 |

*Evaluable patients are those with interpretable biopsies and known best response/clinical benefit data
**One patient (complete responder) did not have quantifiable CD8 staining.
Study ID 45 is PARPi-naïve, whereas all other patients received prior PARPi.
Abbreviations: CR, complete response; PD, progression of disease; PR, partial response; SD; stable disease.

**Table S7. Transcriptomic profiling of each patient (Related to Fig. S4, please see Excel file).**

**Table S8. Differential gene expression of clinical benefit vs. no clinical benefit (Related to Fig. 5, please see Excel file).**

**Table S9. Differential gene expression of clinical benefit vs. no clinical benefit across fork stabilization and replication stress-related genes (Related to Fig. 5, please see Excel file).**

**Table S10. Copy number alteration from whole exome sequencing (Related to Fig. S7, please see Excel file).**
